# Enabling Health Outcomes of Nature-based Interventions: A Systematic Scoping Review

**DOI:** 10.1101/2022.03.16.22272412

**Authors:** Rachel Nejade, Daniel Grace, Leigh R. Bowman

## Abstract

**Background:** The burden of poor mental health and non-communicable disease is increasing, and some practitioners are turning to nature to provide the solution. Nature-based interventions could offer cost-effective solutions that benefit both human health and the environment by reconnecting individuals with nature. Importantly, the relative success of these interventions depends upon the accessibility of green and blue spaces, and the way in which people engage with them.

**Aims and Objectives:** A scoping review was conducted to establish the evidence base for nature-based interventions as a treatment for poor mental and physical health, and to assess whether and how enablers influence engagement with natural outdoor environments.

**Methods:** This scoping review followed the PRISMA-ScR and the associated Cochrane guidelines for scoping reviews. A literature search was performed across five databases and the grey literature, and articles were selected based on key inclusion and exclusion criteria. Exposure was the active engagement with natural environments. The primary outcome was mental health and the secondary outcome was physical health, both defined using established metrics. All data was extracted to a charting table and reported as a narrative synthesis.

**Results:** The final analysis included thirty-nine studies. Most of these focused on green spaces, with only five dedicated to blue spaces. Six nature-based health intervention types were identified: (i) educational interventions, (ii) physical activity in nature, (iii) wilderness therapy, (iv) leisure activities, (v) gardening and (vi) changes to the built environment. Of the 39 studies, 92.2% demonstrated consistent improvements across health outcomes when individuals engaged with natural outdoor environments (NOEs). Furthermore, of 153 enablers that were found to influence engagement, 78% facilitated engagement while 22% reduced engagement. Aspects such as the sense of wilderness, accessibility, opportunities for physical activity and the absence of noise/ air pollution all increased engagement.

**Conclusion:** Further research is still needed to establish the magnitude and relative effect of nature-based interventions, as well as to quantify the compounding effect of enablers on mental and physical health. This must be accompanied by a global improvement in study design. Nevertheless, this review has documented the increasing body of heterogeneous evidence in support of NBIs as effective tools to improve mental, physical, and cognitive health outcomes. Enablers that facilitate greater engagement with natural outdoor environments, such as improved biodiversity, sense of wilderness and accessibility, as well as opportunities for physical activity and an absence of pollution, will likely improve the impact of nature-based interventions and further reduce public health inequalities.

## Introduction

It is estimated that 10% of the global population live with a diagnosed mental health disorder leading to negative health and economic impacts for both individuals and broader society [1]. Of those affected, 10-20% are children – half of whom are already suffering by the age of 14 [2, 3]. Neuropsychiatric and developmental disorders such as ADHD and ASD are particularly common [4], while depression and anxiety are more prevalent among adults [1]. As individuals age into retirement, the risk of mental ill health increases, in part due to social exclusion, loneliness, changes to physical health and the passing of friends and relatives [5]. If population estimates are correct, the global fraction of those >60yrs will have increased from 12% to 22% by 2050 [5]. Therefore in the absence of effective interventions, the global burden of poor mental health will continue to climb.

In financial terms, the combined direct and indirect cost of mental health disorders across the UK in 2013 was estimated at between £70-100 billion annually [6]. Within the European Union (EU), these costs were estimated to be around €798 billion each year [7]. Worldwide, governments and international agencies such as the World Health Organisation (WHO) have responded to the mental health epidemic by increasing funding into mental health research and services [8, 9], yet despite this, first-line treatment for conditions such as depression, ADHD and Generalised Anxiety Disorder (GAD) still rely heavily on medications and approaches such as cognitive behavioural therapy (CBT) [10, 11]. Although these strategies are often effective, medications come with a long list of potential side effects [12, 13], not to mention the financial barriers to access [14, 15]; plus, there are often shortages of skilled mental health practitioners to match the demand for long-term individualised CBT.

In contrast to medicated interventions, there has been renewed interest in “natural” therapies, which are seen as less intrusive and more cost-effective [16]. Meditation, lifestyle changes such as increased physical exercise, community-based activities and engagement with natural environments are emerging as potential alternatives to complement or replace other forms of treatment [16–18]. Indeed, there is growing evidence to suggest that nature-based health interventions (NBIs) can improve mental and physical health outcomes while also addressing the growing patient demand for less intrusive and more cost-effective treatments [16, 19]. That said, challenges exist; NBIs must take place in natural outdoor environments (NOEs) - defined as “any environment in which green vegetation or blue water resources can be found” – and access is becoming increasingly difficult [20, 21]. Indeed there are many geographical, financial and cultural barriers that affect the way we interact with NOEs, and sadly, without significant changes to the way humans live, it is likely that these factors will be compounded by increasing migration away from wild spaces, and further concentration of human populations within urban areas - indeed 68% of the world’s population is expected to reside in urban areas by 2050 [22].

Accordingly, this scoping review attempts to set a baseline for the impact of NBIs on mental and physical health outcomes, as well as better understand the enablers that magnify or diminish engagement with natural outdoor environments.

### Aim

This review aimed to collate and assess the evidence base for NBIs, and to define and assess the effect of enablers on engagement with natural outdoor environments. More specifically, to:

1. locate and review the evidence base for nature-based interventions for mental and physical health outcomes;
2. identify the enablers of, and barriers to, engagement with natural outdoor environments;
3. understand whether these enablers and barriers impact the effectiveness of nature-based interventions on mental and physical health outcomes.

## Methods

### Study Design

This scoping review was conducted according to the Preferred Reporting Items for Systematic Reviews and Meta-Analyses Extension for Scoping Reviews (PRISMA-ScR) guidelines, and the Cochrane guidelines for scoping reviews [23, 24]. A scoping review was considered the most appropriate method to answer the research question, due to capacity to answer broad questions and summarise findings from a heterogeneous body of knowledge [25].

### Study Protocol

The protocol for this scoping review was drafted using the Preferred Reporting Items for Systematic Reviews and Meta-analysis Protocols (PRISMAP) and was revised by the academic team [26]. It was disseminated through MedRxiv, the preprint server for health sciences on the 4^th^ of July, 2020 [19].

### Search Strategy

The search includes terms relating to NBIs: a) ‘green care’, b) ‘blue care’, c) mental health, d) physical health, e) environmental determinants of NOE use and f) socio-economic determinants of NOE use. The primary outcome of interest was mental health, defined using a number of key metrics. The secondary outcome was physical health, based on a number of physiological variables [27]. Several NBI studies have used physical health measures either as their main outcome (e.g. obesity), or as an objective measure to confirm mental health outcomes obtained from self-reporting (e.g. the link between stress and cortisol) [28–50]. All the key words used for the literature search can be found in Supporting Information 1 (S1_Fig 1).

The terminology used in the literature search for green and blue care reflects the varied positions held by researchers and the lack of consensus surrounding their application.

The search terms were used to identify studies from the following five databases: PubMed, The Cochrane Library, Web of Science, Scopus, and OVID (including Embase, PsycINFO, Global Health, MEDLINE, Health Management Information Consortium (HMIC), Transport Database). All search terms were grouped using the Boolean “OR” and were then all combined using the Boolean “AND”, to produce the final number of relevant studies identified by each database. “Snowballing” – or the search of reference lists from included articles – was also performed. To limit the effect of publication bias, grey literature was searched through Google Scholar, governmental and institutional websites (e.g. Public Health England – PHE). Mendeley and Covidence software were used to store, organise, and manage all references. To promote transparency and reproducibility, the full search strategy used for the database PubMed is available in S1_Table1.

### Study Selection Criteria

The process of study selection was done based on the pre-defined inclusion and exclusion criteria and conducted in two stages: 1) title and abstract screening, and 2) full-text screening. Consensus among the three researchers was reached where a study was deemed to sit on the border between inclusion and exclusion. Backtracking of existing reviews was done so that any study included in this scoping review also found in selected reviews was removed from analysis. Duplicates were removed from the search before screening of the articles.

As this is an emerging field, the inclusion criteria for this scoping review were kept intentionally broad. Included were human studies and peer-reviewed articles on green spaces and blue spaces, with physical or mental health outcomes. Any study design was accepted. NOE exposure was based on participants’ presence in nature, whether that was confirmed through participant’s observation, interviews in nature, or through an intervention using activities in NOEs. Any review including at least one study for which NOE exposure was confirmed by these means was included in this review.

Considering the contemporary topic of this scoping review, the search included all results from 1980 onwards. Studies written in both English and French were included. Any studies or reviews not pertaining to health, green spaces and blue spaces, or that were solely descriptive in nature (e.g. commentaries) were excluded. Additionally, studies that only defined NOE exposure based on geospatial indicators were excluded (e.g. Normalized Difference Vegetation Index (NDVI)). To avoid complexities associated with recall bias, we excluded any study that used self-reported measures of engagement with nature (e.g. “number of visits to parks in the last week”) [51, 52]. However, this restriction was not applied to our main outcomes when these were found in studies using self-reporting scores such as GAD-7 and GHQ, as the validity of these measures to assess mental or physical health outcomes has been widely accepted within the scientific community. Additionally, this exclusion criterion would also have greatly reduced the number of available studies [28–50]. The full inclusion and exclusion criteria can be found in Table 1.

**Table 1.** Inclusion and exclusion criteria.

| Inclusion Criteria | Exclusion Criteria |
| --- | --- |
| ▪ All studies and reviews that reported green and blue space exposure (passive or active *interaction* with nature) and physical or mental health outcome of interest, including:<br>· Peer-reviewed articles (e.g. original research and review articles)<br>· Conference proceedings, dissertations/theses and abstracts published in peer-reviewed journals<br>· Grey literature: Google Scholar, governmental, institutional websites, and local health authority publications<br>▪ Published from year 1980 onwards<br>▪ Any study designs and any study written in either English or French | ▪ All studies that reported only NOE exposure without physical or mental health outcomes.<br>▪ All studies that reported NOE exposure only using aggregated data (e.g. NDVI) or using self-reports.<br>▪ Literature including:<br>· Books, book chapters and book reviews<br>· Websites and newspaper articles<br>· Social media content<br>· Commentaries, protocols, and opinion papers<br>· Data not in public domain or unpublished<br>▪ Published before 1980 and in any other language than English or French |

### Data Extraction and Analysis

Data extraction (or charting) was performed using a standardised data extraction form, adapted for this scoping review to address the research questions and objectives (S1_Table2). Content analysis was used to group findings in categories sharing similarities to create a narrative synthesis of the existing evidence informed by the data charting process.

## Results

### Study Selection

The results of the literature search across the five databases and the grey literature were reported using a PRISMA flow diagram (Fig 1). From the original 952 articles, 824 unique studies were identified for title and abstract screening, after the removal of 128 duplicates. Through title and abstract screening, 352 full-text articles were selected and downloaded for a full-text review (i.e. eliminating 472 studies). 313 studies failed to meet the inclusion criteria at full-text screening (reasons detailed in S1_Fig2). A total of 39 articles were selected for the final analysis of this scoping review [53–91].

**Fig 1.**
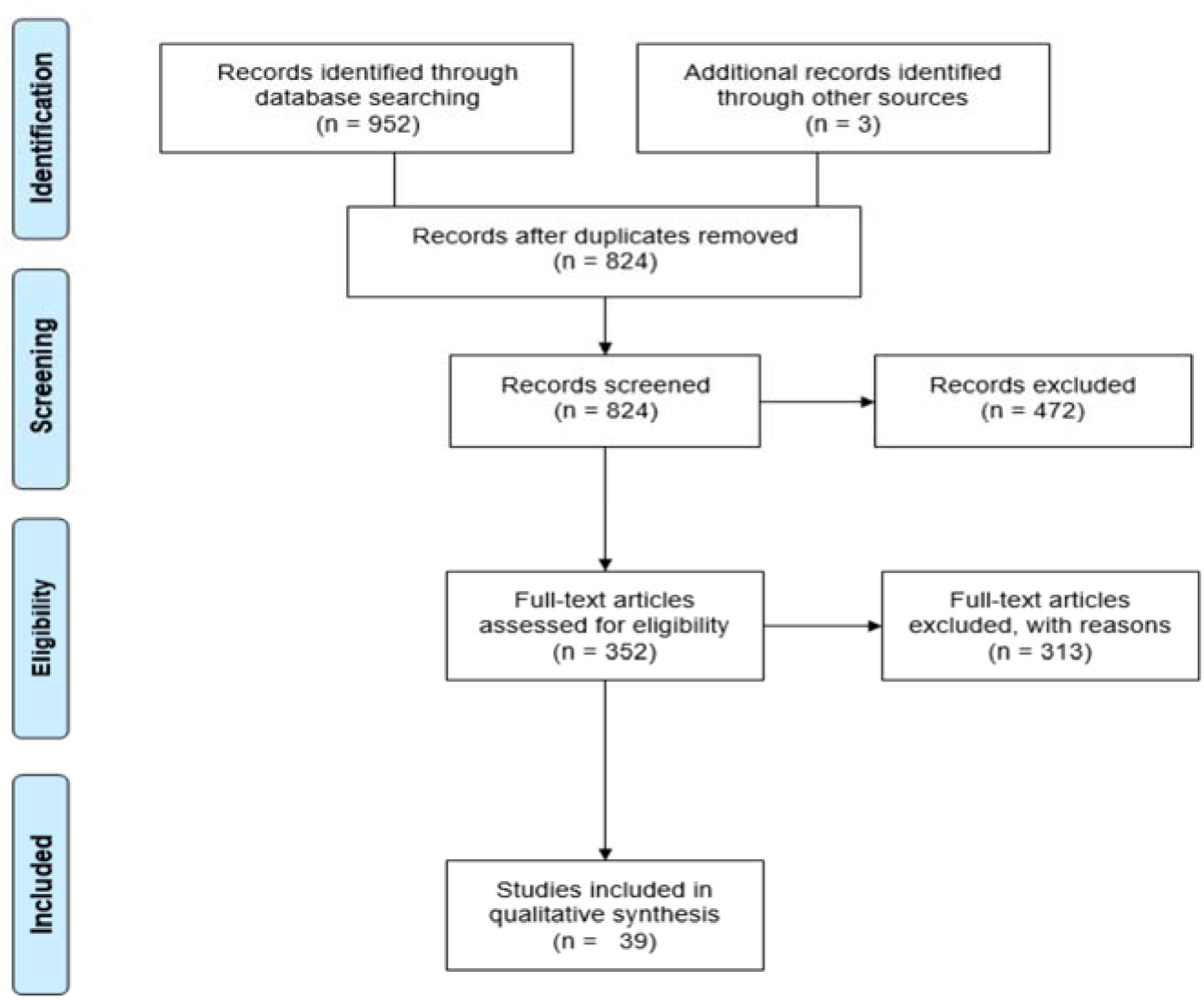
PRISMA flow diagram.

### Descriptive Characteristics

A summary of each charted study can be found in S1_Table3. A total of thirty-nine studies were included in the final analysis, among which eleven were observational, where seven used qualitative methods [55-57, 60, 74, 83, 84], three used quantitative methods [63, 85, 88] and only one used mixed methods [75]. Among the fourteen interventional studies, only one used qualitative methods [66], nine used quantitative methods [58, 59, 61, 62, 64, 65, 67, 81, 91], and four used mixed methods [53, 54, 87, 89]. Finally, among the remaining fourteen reviews, ten included systematic reviews [70-72, 76-79, 82, 86, 90], one was a scoping review [80] and three were literature reviews [68, 69, 73]. All studies were written in English, except for one that was written in French [82]. Additionally, all studies were carried out in the past five years, with the oldest study dating back to 2015 [57].

Overall, the majority of studies (85%) were conducted in higher-income countries (defined using the World Bank classification based on countries’ Gross National Income (GNI) per capita) [92]. Few studies were conducted in upper middle-income countries, including one observational study in Mexico [88], two interventional studies from China [67] and South Africa [61], as well as three reviews including Chinese [76, 82] and Bulgarian studies [76, 80] (Fig 2).

**Fig 2.**
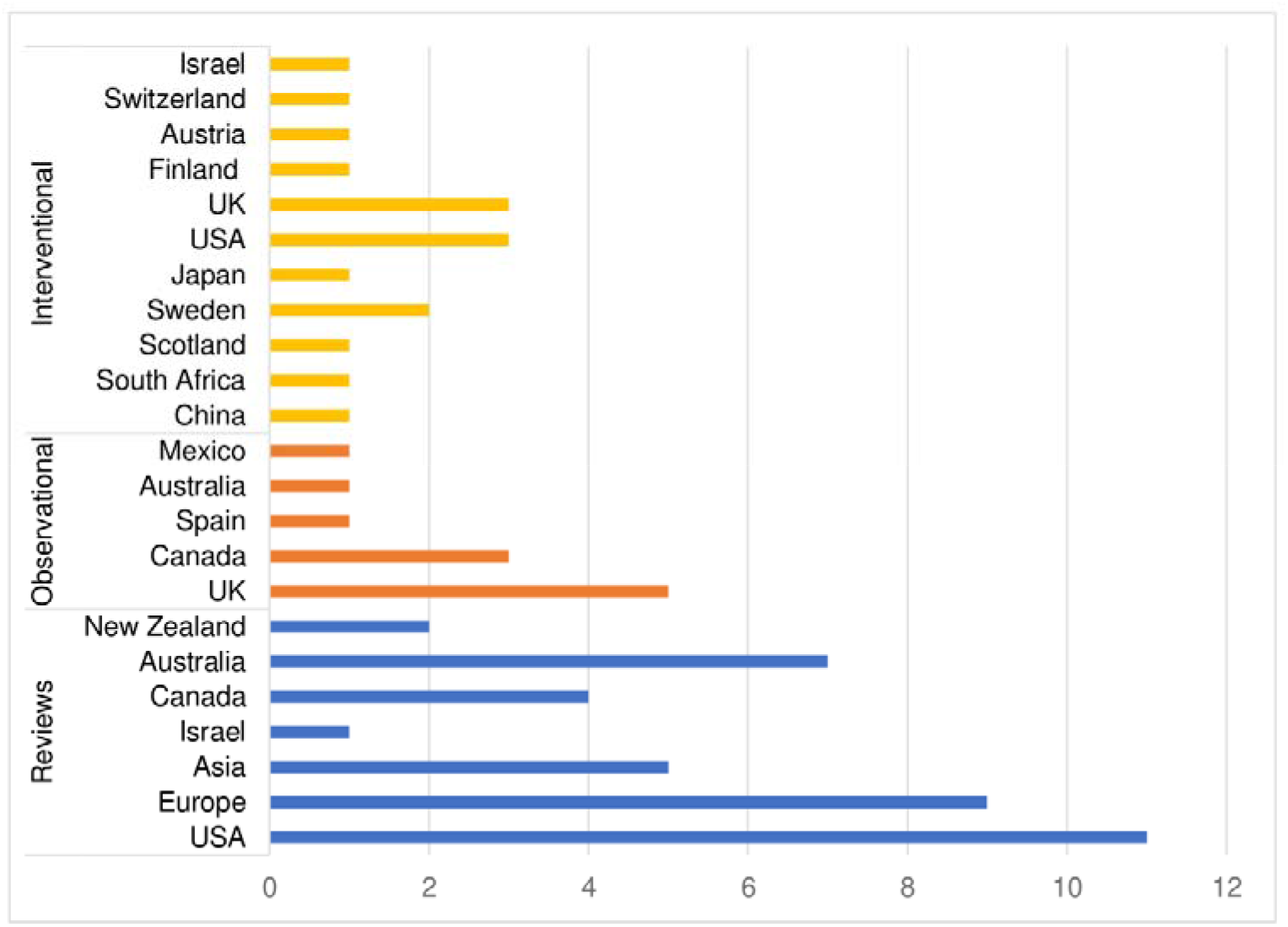
Bar chart depicting the countries included in reviews, interventional and observational studies, grouped by study design.

Twenty out of the thirty-nine studies (51%) assessed the effects of engagement with NOEs on mental and physical health across all age groups, with only ten studies (26%) focusing specifically on adults (18-60 years) [54-56, 58, 59, 62, 63, 67, 81, 91], four (10%) on the elderly (age 60+) [57, 73, 74, 83], as well as four (10%) on children [53, 66, 72, 88] and one (3%) on adolescents (11 to 18 years) [61].

Overall, eight studies (20%) selected participants based on age group [53, 57, 61, 72, 74, 76, 86, 88], two (5%) based on sex (in favour of women) [62, 91], and six (15%) from volunteering [54, 59, 63, 67, 81, 84]. Local residents were recruited in another four studies (10%) [58, 60, 65, 75]. Moreover, eight studies (20%) included patient populations with pre-existing conditions [90]. These looked at people with autism [66], neurological disabilities [73, 78], mental disorders [75, 84, 87] or those undergoing stroke rehabilitation [64]. Noteworthy are the studies that selected participants based on their existing use of natural environments, such as regular swimmers or members of outdoor associations in blue spaces [55, 56, 83], or through involvement in the conservation of green spaces [89]. Finally, eight reviews (20%) did not specify any sample populations [68-71, 77, 79, 80, 82].

### Taxonomy for NOEs

Overall, there were three types of NOEs identified across all studies: (i) green spaces: 51% (n=20), (ii) blue spaces: 13% (n=5), and (iii) a mix of both 36% (n=14).

Green spaces encompassed both urban and rural environments, and most studies described green spaces as urban parks [57, 62, 65, 69, 74, 82, 85, 88, 91], natural environments [63, 68, 70, 72, 86] urban forests [53, 62, 78, 81] or as gardens [64, 73, 74, 78]. Other areas or features of green spaces were used less often, such as farms [53, 66, 78], micro-features of green spaces [57, 74], national parks or reserves [60, 89], a game reserve [61], urban stream corridor [55], playground [72], meadows [54], bog [89], or neighbourhood greenness [77]. Similarly, blue spaces also covered urban and rural environments and were characterised by the terms: sea [56, 90], blue environments [70, 86], river [53], fountain/ seawall [74], coastal area [59], loch [61], wetlands [87], wilderness [90], ocean and beaches [83]. Finally, grey areas were typically considered as urban environments: urban city [54, 62, 65, 91], built environment [58, 79], urban sidewalk [59], shopping mall [62], hospital [64], urban landscape [72], roadside [81], home [91], swimming pools [83] and a field near a housing development [89].

### Nature-Based Health Interventions (NBIs)

All NBIs and their related activities reported across the selected studies were categorised, as illustrated in Fig 3. Six types of NBI were identified: (i) educational intervention, (ii) physical activity, (iii) wilderness therapy, (iv) leisure activity, (v) gardening, and (vi) changes to the built environment.

**Fig 3.**
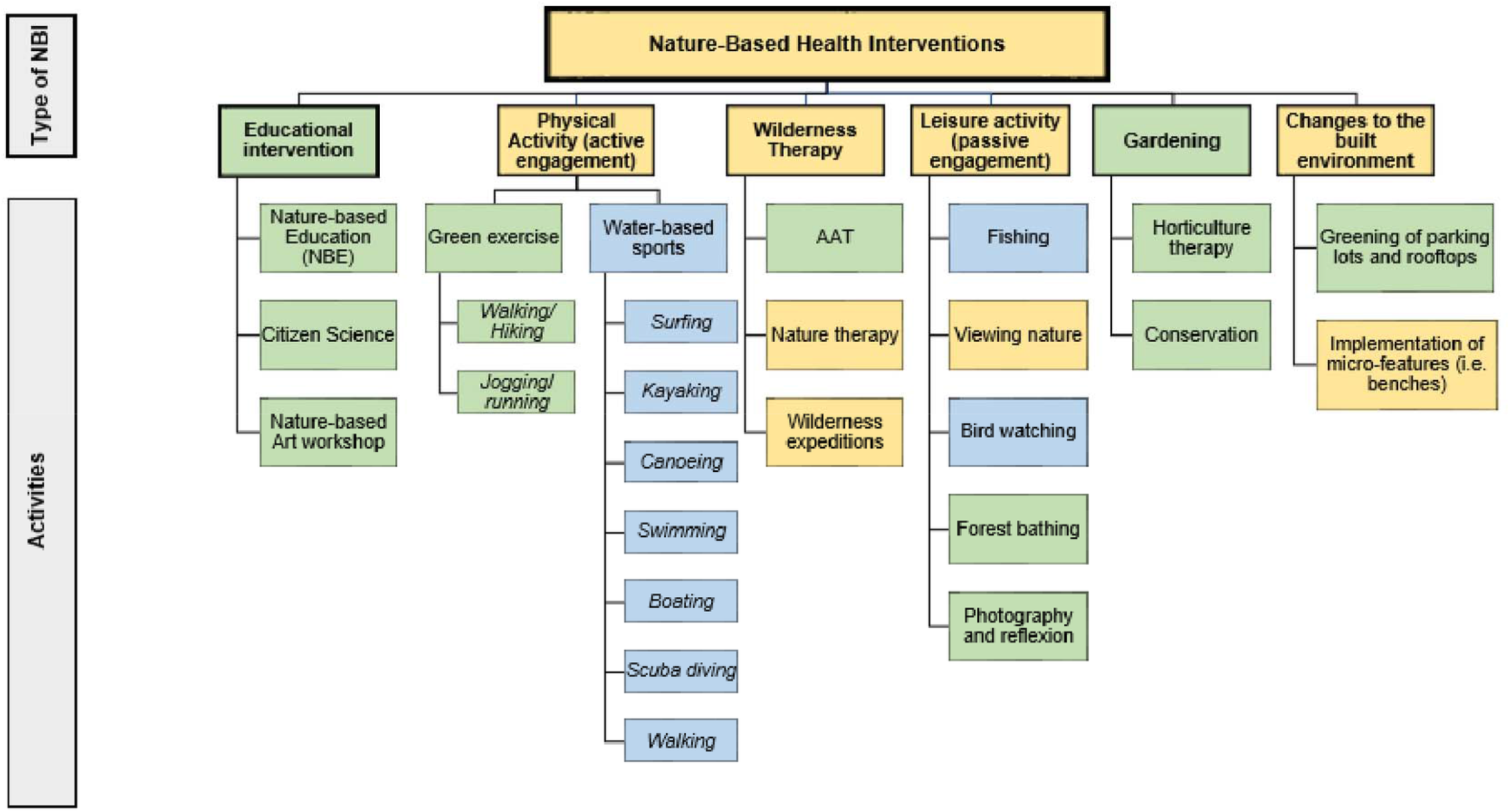
All types of nature-based health interventions found in the selected studies. Green=green spaces, blue=blue spaces and yellow=both green and blues spaces.

### Health Outcomes and Nature-based Interventions

All reported outcomes and their associated enablers are listed in S1_Table4. Almost all of the studies included at least one psychological health outcome [53, 54, 56–66, 68–87, 89–91], except for three that focused solely on physical activity [55, 88] and cardiovascular outcomes [67]. Many studies used multiple outcomes, and each of these is reviewed and discussed in the following order: (i) mental health outcomes, (ii) physical health outcomes, (iii) physiological outcomes, and (iv) cognitive health outcomes.

Overall, there are clear positive trends between NOE engagement (through voluntary participation or primary care intervention) and psychological, physical and cognitive health outcomes. These data are described in Fig 4 by the bars labelled “positive findings”. Note that in applicable studies [56, 73–75, 83, 87], a decrease in the measurable outcome was considered a “positive finding” where this resulted in a gain for the individual e.g., a reduction in social isolation. The studies displayed as “negative findings” refers to studies where health outcomes led to mixed or no positive effects [59, 70, 71, 76, 81, 82, 87, 91].

**Fig 4.**
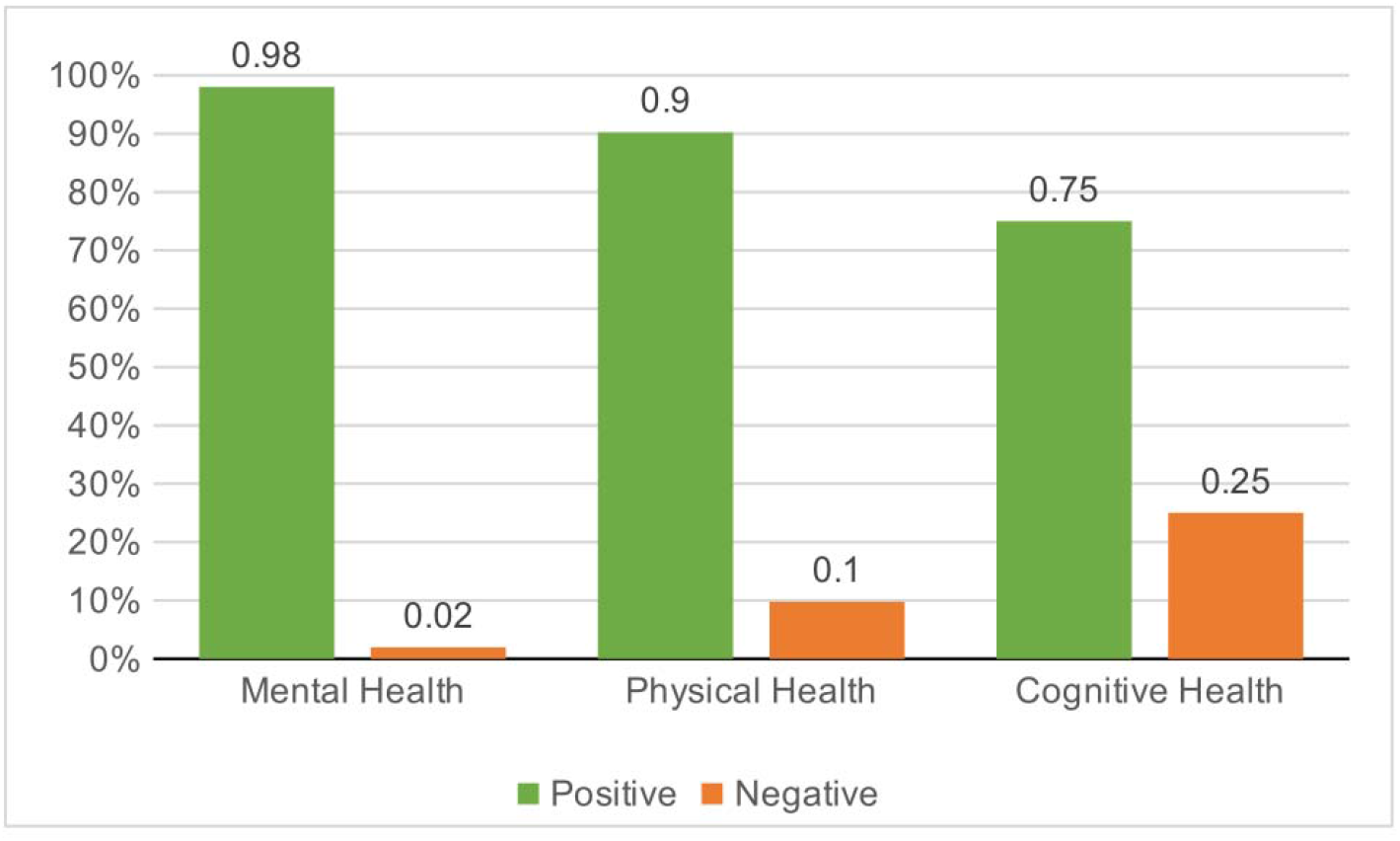
Proportion of positive and negative findings stratified by health outcomes.

### Mental Health Outcomes

Mental health was the most commonly studied outcome (62%) and was further divided into four categories: (i) psychological health, (ii) emotional health, (iii) social health and (iv) stress. There were improvements across all mental health outcomes when engaging with nature (98%), with only one study reporting no effect (2%) [71]. No negative effects were found.

### Psychological Health

Overall, engagement with NOEs led to improvements in psychological health outcomes (63%), and reduced depression and anxiety scores (31%). One study reported no differences on wellbeing measures (6%) [71].

Engagement with NOEs led to an improved quality of life as assessed by measures of HRQoL [53, 64] or QoL [69, 86]. Only one study reported improved “perceived mental health” (e.g. restoration) of stream-corridor users, assessed using qualitative interviews [57]. Regarding measures of wellbeing, these were the most studied outcome for psychological health (50%) and was usually positively associated with NOE engagement. It was measured differently across studies and relied on measures of hedonic and eudaimonic wellbeing [71], perceived wellbeing [56, 73–75, 83, 90] and general wellbeing [54, 58, 63, 68, 71, 76, 86, 87]. Only one systematic review reported mixed effects, which the authors attributed to poor study design and quality [71]. Finally, measures of depression [63, 65, 78, 80] and anxiety [64, 65, 78, 81, 87] decreased when engaging with NOEs.

### Emotional Health

There was a positive effect of NOE engagement on emotional health outcomes across all studies. Most reported improved affect [58, 64, 70, 73, 81–83, 86, 87], mood [62, 65, 79, 80, 89, 91], self-esteem [61, 73, 80, 84, 90], self-confidence [75] and vitality [79]. Others reported decreases in negative affect [63, 81, 83, 86], mood disturbances [65], agitation [73, 78] and in behavioural problems (e.g. hyperactivity or violence) [72, 73, 80, 82], which all translated into improved emotional health.

### Social Health

Overall, engagement in NOEs led to improved social health across 100% of the fourteen studies that assessed their effects. Six studies reported reduced social isolation [56, 73–75, 83, 87] and one found reduced social discomfort [91] following engagement in natural environments; while seven noted increased social connectedness between individuals [66, 68, 78, 82–84, 90].

### Stress

Several studies assessed the effects of engagement with NOEs on stress. 100% of studies reported positive associations with psychological resistance [54, 56, 90], perceived restoration [59, 60, 62, 65, 82, 91] and stress reduction [54, 63, 66, 73, 81–84, 89]. Only one study found a decrease in psychological distress [80], and three other studies found decreases in perceived stress [63, 86, 87], which all translated into health benefits.

### Physical Health

89% of studies that looked at physical health measures in relation to engagement with NOEs found benefits across a range of outcomes, except for 11% that yielded no effects for measures of obesity.

### Physical Activity

All measures of physical activity in natural environments demonstrated that engaging in NOEs led to increased physical activity. This was measured in several ways. Some studies used measured of leisure-time physical activity [55] or reported use after urban green spaces interventions [69, 74, 82]. Others focused on increased exertion post-engagement with NOEs, using measures of moderate to vigorous physical activity [79, 88].

Increased use of NOEs for various activities like swimming [56] or walking in nature [60, 76], using measures of perceived physical activity [56, 57, 68] and physical fitness [90] as another way to reach these conclusions, as well as using decreased sedentary time, in children populations [88].

### Sleep and Recovery

One systematic review assessed the effect of engagement with green spaces on sleep during a walking intervention and found that engagement led to improvements in sleep quality and quantity [77]. Similarly, one study reported improved recovery from mental disorders after engaging in therapeutic horticulture as part of a recovery program [75].

### Motor Functioning

Motor functioning was assessed differently by two studies [64, 74]. Ottoni et al. (2016) reported improved mobility after walking interventions in green spaces (74) compared with data from Pálsdóttir et al. (2020) that reported reductions in disability after engaging in horticulture therapy for post-stroke patients [64]. Overall, improvements in disability were reported, in both intervention and control groups, suggesting that the therapy itself may facilitate recovery more than the type of environment [64].

### Overall Physical Health and Fatigue

100% of those studies measuring physical health outcomes found a positive association between physical health and NOE engagement when measured by General Health Questionnaires (GHQ) [72, 80, 87]. Post-stroke fatigue (or PSF) was used by Pálsdóttir et al. (2020) as their main outcome and decreased following horticulture therapy [64]. Importantly, both the intervention and control group experienced decreases in PSF, thereby reducing the importance of the intervention in this context over other mainstream standards of care.

### Obesity and Mortality

Four studies reported little to no effects on obesity (measured using BMI) after engagement with NOEs [70, 76, 82, 88]. Regarding mortality, only two studies investigated how NOE engagement affected all-cause mortality [70, 79]. Both studies found a decrease in mortality following changes to the built environment [70] and after engaging in physical activity in nature [79].

### Physiological Health

Although physiological measures (8%) were often preferred over subjective measures of health, only seven studies used physiological measures of cardiovascular health (86%) [54, 62, 65, 67, 79, 91] and only two focused on stress (14%) [62, 82].

### Cardiovascular Health

Cardiovascular health was measured using blood pressure (BP) [62, 65, 67], baseline resting heart rate [54, 65, 67, 69] and heart rate variability [62, 79, 91]. Heart rate was found to decrease in 80% of studies looking at this measure, except for one [65]. Similarly, BP was found to decrease in three studies except for one by Ana et al. (2019) which found no changes in BP after forest bathing [67]. Results were inconclusive as well for heart rate variability, which tended to increase after NOEs exposure in two studies [62, 91], but had no effects in another [79].

### Stress

Physiological measures of stress were determined using cortisol samples, and in two studies, there was a decrease in cortisol levels after engaging in NOEs [62, 82].

### Cognitive Health

Although not initially included, cognitive health outcomes were identified on several occasions (7%) during the analytical process and were therefore considered important for this review. Overall, NOE engagement had positive effects on cognitive health, by improving cognitive functioning and by reducing ADHD symptoms. Findings on memory were inconclusive.

### Cognitive Functioning

86% of studies focusing on cognitive functioning were positively associated with NOE engagement, except for one study which reported no change in attention retention [59]. However, attention retention was improved after exposure to natural environments in three studies [66, 79, 82], along with attention restoration [54, 72]. One study also showed an improvement in children’s Science, Technology, Engineering, and Math (STEM)-capacity following NBE intervention [53].

### Memory

Memory was only assessed in four studies and yielded mixed findings. Indeed, whereas one found a positive association between spatial working memory and engaging in NOEs [72], the other found no effects [91]. Similarly, for executive functioning, one study found no effects [81], while the other saw improvements in executive memory [78].

### Symptom Reduction

Trained therapists during a wilderness expedition noticed a decrease in ADHD symptoms for children living with autism after exposure to animals and the natural environment [66] – which was supported by McCormick R. (2017) in her systematic review [72].

### Engagement with Natural Outdoor Environments: Barriers and Enablers

Overall, there were several types of enablers identified throughout the selected studies. These included environmental, social, individual, and structural processes, along with opportunities for physical activity and stress reduction. Poor study design and quality were considered barriers across all studies. A description of each enablers’ category can be found in S1_Fig3.

Modulators were divided into those that facilitated engagement: enablers (∼78%) versus those that hindered engagement: barriers: (22%). A summary of all enablers and barriers stratified by health category is shown in Fig 5.

**Fig 5.**
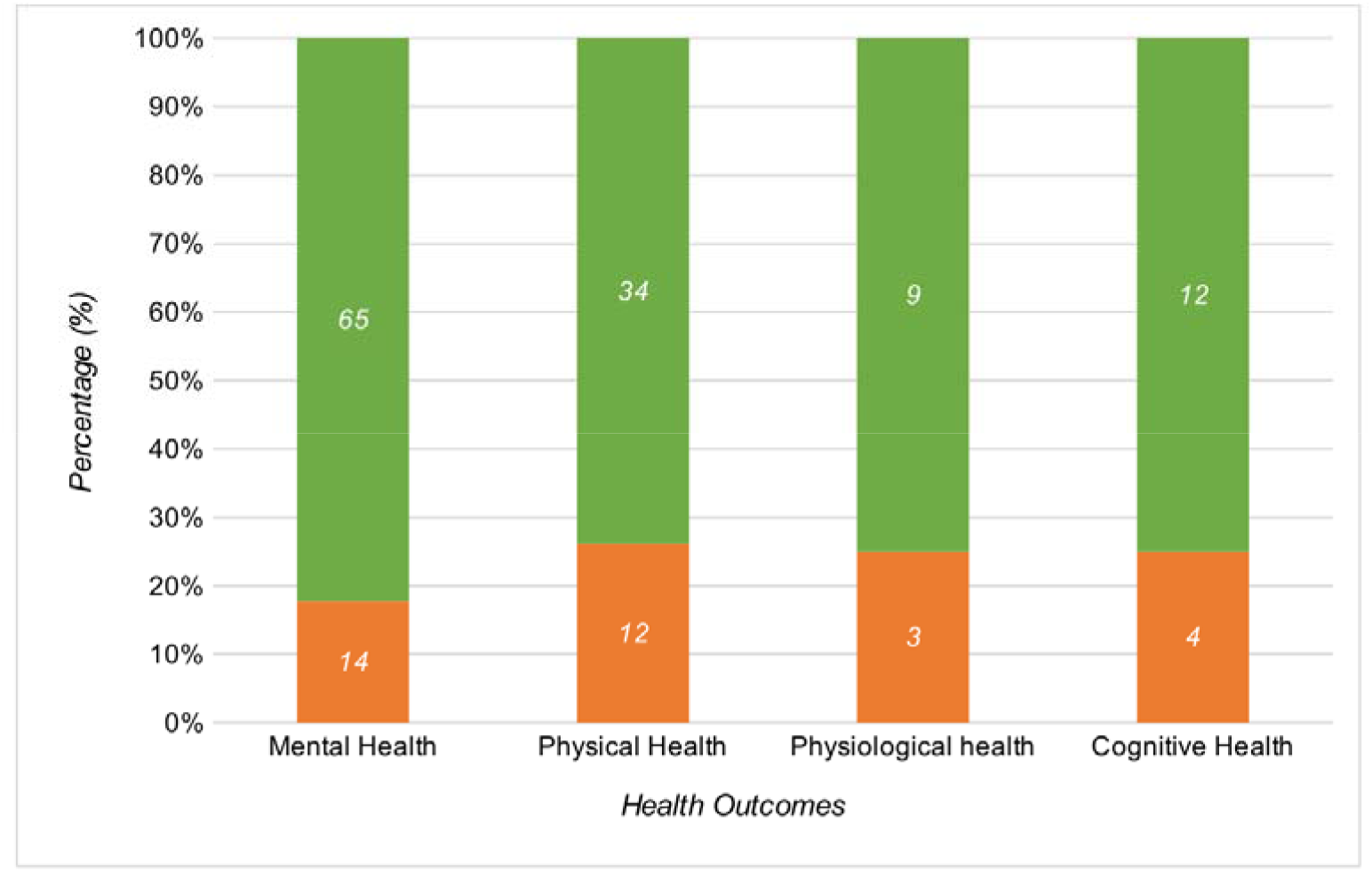
Number and proportion of enablers and barriers for each health outcome. Enablers in green; barriers in orange.

### Environmental Processes

Most facilitators focused on environmental processes (38%), the most common being the type of environment (66%), where natural environments facilitated health benefits over built environments (i.e. swimming pools, city centres, shopping mall, etc.) [53-56, 58, 59, 62, 63, 65-67, 70, 72, 73, 77-84, 86-91]. Although the variety of green spaces and blue spaces descriptors makes comparison between studies difficult, some studies have found that urban forests were better than urban parks as they reduced cortisol levels [62], blood pressure [54, 62, 65] and heart rate [62, 67, 79], while increasing heart rate variability [62, 91]. Interestingly, one study even found that heart rate benefits were amplified if that forest was made of maple trees as opposed to birch or oak trees, while blood pressure would not change if the temperature, humidity and light spectrum (i.e. green/blue light ratio) were too high [67]. Similarly, users of blue spaces preferred wilder and more available water environments (e.g. ocean) as they amplified psychological health benefits, through increases in wellbeing, as well as social health benefits by reducing social isolation [56, 83]. Typically, biodiversity was shown to facilitate wellbeing [87], psychological restoration [85], social connectedness [87], positive affect [83] and overall health [87], while reducing anxiety [87] and stress [87]. It is important to note that biodiversity may also present a barrier if perceived as threatening or harmful. The presence of sharks in blue spaces is one such example of this [83]. Other environmental processes, such as good weather [83], heat reduction [70], seasons [65], perceived aesthetics [68], nature connectedness [58, 71], the presence of farm animals for autistic children [66] as well as sensory qualities of the environment (i.e. sound) [59, 60] were all found to also improve mental, physical, physiological and cognitive health, however other detrimental environmental processes, such as air and noise-related pollution [62, 76], negated these positive effects.

### Structural Processes

Structural processes were the second most common enablers discussed in this scoping review (37%). Among them, good accessibility was most commonly reported (24%), as it facilitated improvements in perceived mental health [57], overall health [72], positive affect [70], physical activity in NOEs [55, 57, 74, 82] and in attention restoration [72], as well as reducing social isolation [74], motor disability [74], behavioural problems and psychological distress [80]. Similarly, geographic proximity to NOEs was also mentioned several times (11%) as facilitating wellbeing [76], physical activity [60, 69, 76], cognitive functioning and spatial working memory [72].

The type of intervention was also reported by six studies (16%) as facilitating the health benefits gained from engaging in NOEs. Britton et al. (2018) and Ottoni et al. (2016) recognise that activities in blue spaces, such as surfing or swimming, contribute to rehabilitation, stress reduction and health promotion [83, 90], and complementary evidence demonstrates that therapeutic horticulture led to improvements in PSF [64] and reductions in agitation for older adults [74]. Additionally, viewing nature decreased blood pressure [62] and improved executive memory [78]. Interestingly, the longer the activity lasted, the better the outcomes [53, 61, 87, 88]. One study found that activities performed in the afternoon instead of the morning, improved sleep quality and quantity – believed to be caused by a two-process model where sleep and waking are regulated by circadian rhythms and homeostasis [77, 93]. Good group organisation, transportation, staff attitudes and knowledge were also considered facilitators of the associations between health and nature [87]. However, when NBIs have limited resources, the strength of these associations is reduced [73, 90], and hence, good NBI quality and design can amplify the health benefits gained from nature.

The quality and design of NOEs were also found to amplify health benefits when engaging with nature, as the presence of micro-features of the environments (e.g. benches) was found on several occasion to improve wellbeing, positive affect and self-esteem, while reducing social isolation and stress in individuals with dementia [73]. Older adults also found that benches could help decrease social isolation [74] and improve their mobility and physical activity in NOEs [74]. Other studies also found general increases in physical activity and positive affect when these features were present [55, 70, 80]. Overall, positive changes to the environment through the implementation of micro-features were found to facilitate engagement in NOEs.

### Individual Processes

Most individual processes across the selected studies were considered barriers (74%) as opposed to facilitators (26%).

Safety concerns were the most common barriers to engaging in NOEs (24%), as they worsened perceived mental health [57], positive affect [70, 73], perceived restoration [60], physical activity [55, 57], wellbeing and self-esteem [73] while increasing social isolation and stress [73]. Stigma was another recurrent barrier found across studies (12%) that diminished perceived wellbeing [73, 90], physical activity [56], physical fitness, social connectedness and psychological resistance [90], as well as positive affect and self-esteem [73], while increasing social isolation [73] and stress [73].

Other barriers such as social prejudice [73], fear [56, 90], negative self-perceptions [57, 73], poor self-confidence [73], individual factors (e.g. time pressure, changing identities) [74, 77] and deprivation [80, 84], were also detected. Conversely, some individual processes were found to facilitate the relationship between nature and health. These included cognitive functioning [72], some intrapersonal processes (i.e. individual preferences) [68], gender – whereby women tended to benefit more than men [61, 74], and age – since younger adults and children had increased health benefits from engaging in NOEs due higher engagement in physical activity than older adults [82].

Lower socio-economic status (SES) and ethnicity were identified as both facilitators and barriers. While one study found that being South Asian living in the UK led to worse health outcomes than being British white [80], another found that Arab women benefited more than Jewish women when engaging in NOEs [91]. The latter was thought to be influenced by levels of comfort at home, where Jewish women reported feeling more comfortable in their home than Arab women did and therefore gained fewer marginal improvements than Arab women when engaging in NOEs [91]. Similarly, lower SES was found to increase the health gains through NOE engagement [82], whereas another found it led to worse health outcomes [76].

### Opportunities for Physical Activity

Opportunities for physical activity were the third most frequent facilitators found across studies (11%). They included physical activity (72%) and active engagement in NOEs (18%) as both were found to magnify the benefits for mental health [56, 58, 63, 71-73, 75, 79–82, 89], physical health [56, 72, 75, 80], physiological health [79] and even cognitive health [72, 78, 79]. However, these benefits would be reduced if participants were injured or had mobility difficulties [74, 78]. Physical activity could therefore be another mechanism by which nature positively influences health.

### Social Processes

Social processes were not as common as other enablers (7%) but were found nonetheless to influence the impact of nature on health. The presence of other people was the most common facilitator (29%) and barrier (29%) across studies considering social processes. Indeed, two studies reported that sharing the experience of engaging in NOEs with others could facilitate gains in physical activity [55], recovery from mental disorders [75], social connectedness, self-esteem, and self-confidence [84], while reducing social isolation [75]. However, if other individuals were perceived as safety risks, wellbeing and physical activity would decrease while stress would increase [63].

Additionally, social interactions, interpersonal processes, group membership and the presence of caregivers also facilitated positive gains in psychological [68, 78, 89], social [68, 83] and physical [68, 89] health. Therefore, social processes are other mechanisms through which health benefits can be gained from nature.

### Opportunities for Stress Reduction

Despite abundant evidence from the literature review, only 1% of all enablers focused on opportunities for stress reduction. Stressful life events were perceived as barriers, as they caused decreased quality of life, wellbeing, positive affect, psychological resistance, and STEM capacity for children [52, 63], while worsening depression in adults [63], however, engaging in NOEs was shown to reduce stress in 100% of all studies looking at stress-related outcomes, considered measures of psychological health in this review [53, 63, 66, 73, 81–84, 89]. Therefore, evidence for stress reduction as a mechanism in the relationship between health and nature is moderate, but not as conclusive as other enablers.

### Study Quality and Design

Methodological choices when conducting studies (9%), such as the study design (44%), study quality (44%) or the choice of measurements (12%) were all found to negate the relationship between health and nature across selected studies [59, 70, 71, 76, 81, 82]. They were responsible for the lack of evidence between NOE engagement and obesity [70, 76, 82], wellbeing [71], heart rate variability [79] and on measures of memory [81] and cognitive functioning [59]. Therefore, the methods used within studies also act as a potential mechanism on nature and health.

## Discussion

The aim of this scoping review was to explore and document the evidence base for nature-based interventions, to identify factors that drive engagement with natural outdoor environments and to examine whether these impact the effectiveness of nature-based interventions. Of the 39 included studies, nature-based interventions were found to have improved physical, mental and cognitive health outcomes across 98%, 90% and 75% of articles respectively (Fig 4). In addition, the evidence suggests that enablers have a significant role in magnifying or diminishing interactions with NOEs -78% facilitated engagement compared with 22% that reduced engagement (Fig 6).

**Fig 6.**
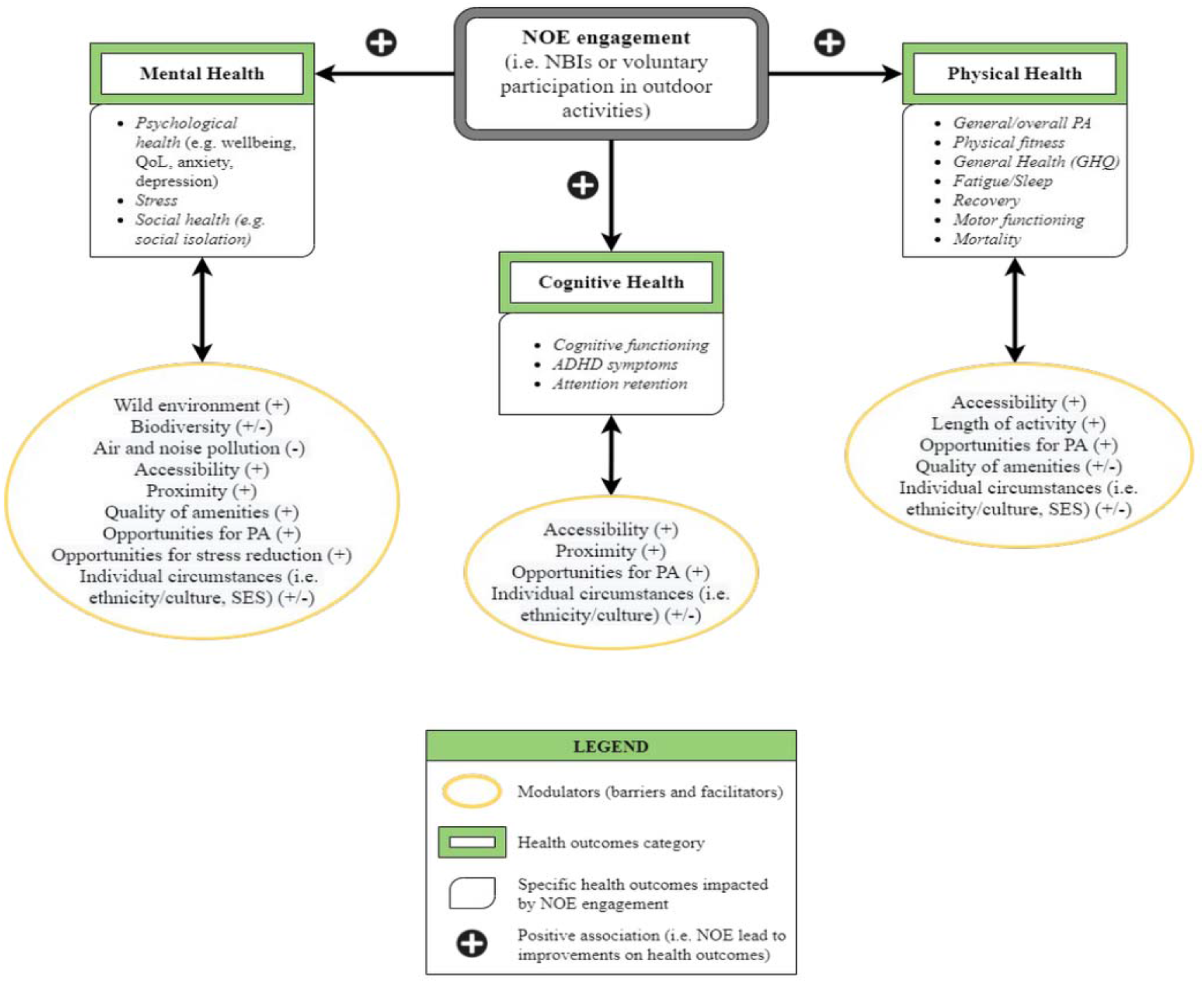
Impact of NOE engagement and enablers on health.

### NBIs and Health Outcomes

As a species, humans have become increasingly sedentary. Offices, schools, homes and public spaces have been designed to optimize and prioritise efficiency. At least in part, this is driving the increase in non-communicable diseases and poor mental health [94]. Moreover as individuals continue to seek work in urban areas - far from wild spaces - the opportunity to interact with green and blues spaces is diminished. Indeed over 50% of people worldwide currently live in urban areas, projected to increase to >68% by 2050 [22, 95].

Considering this, perhaps it is not surprising that the reintroduction of nature into a person’s life, irrespective of baseline physical and mental health characteristics, is often a positive influence [96]. Research shows that, on average, individuals living in urban areas with more green space have both lower mental distress and higher well-being scores [97]. ‘Forest-bathing’ (shinrin-yoku) in Japan has been shown to significantly lower salivary and serum cortisol levels when compared to control groups [98]. And data from Niedermeier et al., 2017, found that hiking resulted in a statistically significant increase in “affective valence” – i.e. pleasure, when compared to a sedentary control group and an indoor exercise group [99]. In this scoping review, 98%, 90% and 75% of articles reported positive outcomes across mental, physical and physiological outcomes respectively, demonstrating a clear link between exposure to NOEs through NBIs to measureable improvements in health. However, study design was a mix of observational, interventional and reviews, indicating that further improvements in study design are required to better quantify the relative effect of NBIs on health outcomes. Yet there are some known theories that might begin to explain the mechanisms behind these results. Indeed it is postulated that when in natural outdoor environments individuals experience a reduction in “rumination” – a maladaptive pattern of self-referential thought that is associated with heightened risk for depression and other mental illnesses [95]. Functional MRI scanning performed on individuals who had spent 90 minutes on a nature walk showed reduced neural activity in the subgenual prefrontal cortex (sgPFC) – an area of the brain that displays increased activity during sadness and rumination. In contrast, participants who went on an urban walk did not show these effects (95).

Such data makes clear the physiological responses that NBIs elicit in humans, and while further granular data are required, the mounting body of evidence is generally in support of nature-based interventions for the prevention and treatment of physical/ mental health ailments. Indeed the science is now starting to inform national healthcare policy via the introduction of ‘green prescriptions’, which are clinically prescribed NBIs used to prevent and/ or treat physical and mental disorders [100, 101].

The broad evidence base uncovered by this scoping review continues to demonstrate the effectiveness of NBIs to improve mental, physical and cognitive health outcomes. Indeed the findings support national policies that integrate NBIs as effective preventative and curative tools for public health [16, 19, 100, 101].

### Enablers and Barriers of NBIs

#### Biodiversity and Wilderness

Our findings on the importance of biodiversity and wilderness as drivers of impactful NOE engagement provide support for a broader interconnectedness between humans and wild spaces. This applies to all projects at any scale, from school expeditions through urban greening to broader rewilding. Enabling interaction with NOEs through accessibility (both geographic proximity and improved infrastructure) magnifies the health benefits of NOEs [55, 57, 60, 69, 70, 72, 74, 76, 80, 82], and at the same time facilitates interaction between the public and natural ecological systems [102].

There exist a variety of example practices, for instance, the creation and maintenance of long-distance trails [102], increasing the sense of ‘wild’ in urban green spaces [83, 85, 87] and a departure from meticulous park management [55, 70, 73, 74, 80]. These approaches result in increased ‘quality’, accessibility and biodiversity, leading to plausible health gains through greater NOE engagement [55, 70, 73, 74, 80, 83, 85, 87, 102]. Indeed this recommendation fits within the broader International Union for Conservation of Nature (IUCN) vision for human interactions and ecosystem health to “[…] protect, sustainably manage, and restore natural or modified ecosystems, that address societal challenges effectively and adaptively, simultaneously providing human well-being and biodiversity benefits” [103].

#### Air and Noise Pollution

Our findings also support wider initiatives targeting reductions in air and noise pollution, as these were found to negatively impact the time that users would spend practicing physical activities in NOEs [62, 76]. Cleaner, greener environments would also encourage physical exercise and contribute to national and global targets to mitigate climate change [62, 76, 104]. Indeed nature-based initiatives, such as de-pollution and re-naturalisation of urban sites, are currently under consideration by the EU Commission as methods to achieve an increase in the number of publicly available green spaces and reverse social inequalities [104, 105].

### Socio-economic Status and Stigma

Cultural and ethnic differences, as well as deprivation, were found to limit the health benefits gained from engagement with NOEs. Minority groups living in more deprived areas, with lower access and quality of green spaces, had more behavioural difficulties than non-minority groups [80, 91]. Despite mixed findings in this review [76, 82], there remain existing inequalities concerning access to urban green infrastructure, as well as inequalities in the exposure to health hazards (e.g. air and noise pollution), particularly for vulnerable groups such as children, the elderly, and individuals of lower socio-economic status [106]. These inequalities are well documented in urban areas across many European countries, and likely exist globally, highlighting the need for urban greening initiatives that work towards reducing social barriers to access, and use of, green and blue environments [106, 107].

### Proximity and Opportunities for Physical Activity

The sedentary lifestyle characterising modern society has also led to a clear reduction in physical activity across age groups [102]. As regular physical activity has been shown to reduce certain health risks such as cardiovascular diseases, or symptoms of depression and anxiety, health agencies such as the WHO have urged governments to promote physical activity to their populations as a way to limit the growing burden of ill health [27, 108].

The results from this review support the need for enhanced engagement in physical activity, and especially when practiced in green or blue environments – as the mental, physical and cognitive benefits gained from such engagement were amplified in these environments. This was also influenced by structural enablers, where good accessibility [55, 57, 74, 82] and geographic proximity [60, 69, 76] led to increased physical activity in NOEs.

This is an important finding for policy makers, as it highlights the need to consider access and proximity to green and blue spaces when designing health interventions promoting physical activity. For example, the establishment of free shuttle services for individuals living further than a pre-determined distance to NOEs could reduce social inequalities of access and use of these environments.

### Limitations

Methodologically, the exclusion of studies based on self-reported measures of exposure (e.g. number of visits in the last month) could have precluded the inclusion of additional relevant studies to this review. However this was deemed necessary to limit the inherent risk of recall bias in these studies, which could have impacted the strength of the results. The absence of critical appraisal of individual sources of evidence precluded the possibility for our results to lead to statistically significant conclusions. Nevertheless, scoping reviews as per PRISMA-ScR guidelines do not necessarily require critical appraisal of the evidence for structural integrity; as a minimum, they promote a stronger evidence-base [23].

The comparison between health outcomes and types of green spaces or blue spaces was made difficult due to the variety of terms used to describe these areas. Similarly, for nature-based interventions, direct quantitative comparisons were difficult due to absence of magnitudes, relative effects, varied heterogeneous study designs and sample sizes.

## Conclusion

Further research is still needed to establish the magnitude and relative effect of nature-based interventions, as well as to quantify the compounding effect of enablers on mental and physical health. This must be accompanied by a global improvement in study design. Nevertheless, this review has documented the increasing body of heterogeneous evidence in support of NBIs as effective tools to improve mental, physical, and cognitive health outcomes. Enablers that facilitate greater engagement with natural outdoor environments, such as improved biodiversity, sense of wilderness and accessibility, as well as opportunities for physical activity and an absence of pollution, will likely improve health outcomes and further reduce public health inequalities.

## Supporting information

PRISMA ScR Checklist

## Data Availability

All data produced in the present work are contained in the manuscript.

## Acknowledgments

Some of the work in this article was adapted from Nejade RM’s Masters in Public Health Thesis: *Reviewing the Evidence for Nature-Based Health Interventions and their Modulators: A Scoping Review*. 2020, Imperial College University.

## Supporting Information

S1_Fig1. Key database search terms.

S1_Table1. Search Strategy in PubMED.

S1_Table2. Data extraction form (for original and review studies).

S1_Fig2. PRSIMA flow diagram, with reasons for exclusion detailed among the 313 excluded studies during full-text screening.

**S1_Table3. Charting table of each individual studies included in this scoping review.** Where cases highlighted in green=GS, blue=BS and yellow=both.

**S1_Table4. All outcomes present in selected studies and categorised through content analysis.** Numbers highlighted green=GS, blue=BS and yellow=both.

S1_Fig3. Description of key modulators to NOE engagement.

