## Supplementary figures and images for "Enabling Health Outcomes of Nature-based Interventions: A Systematic Scoping Review"

### Supplementary Figure 1

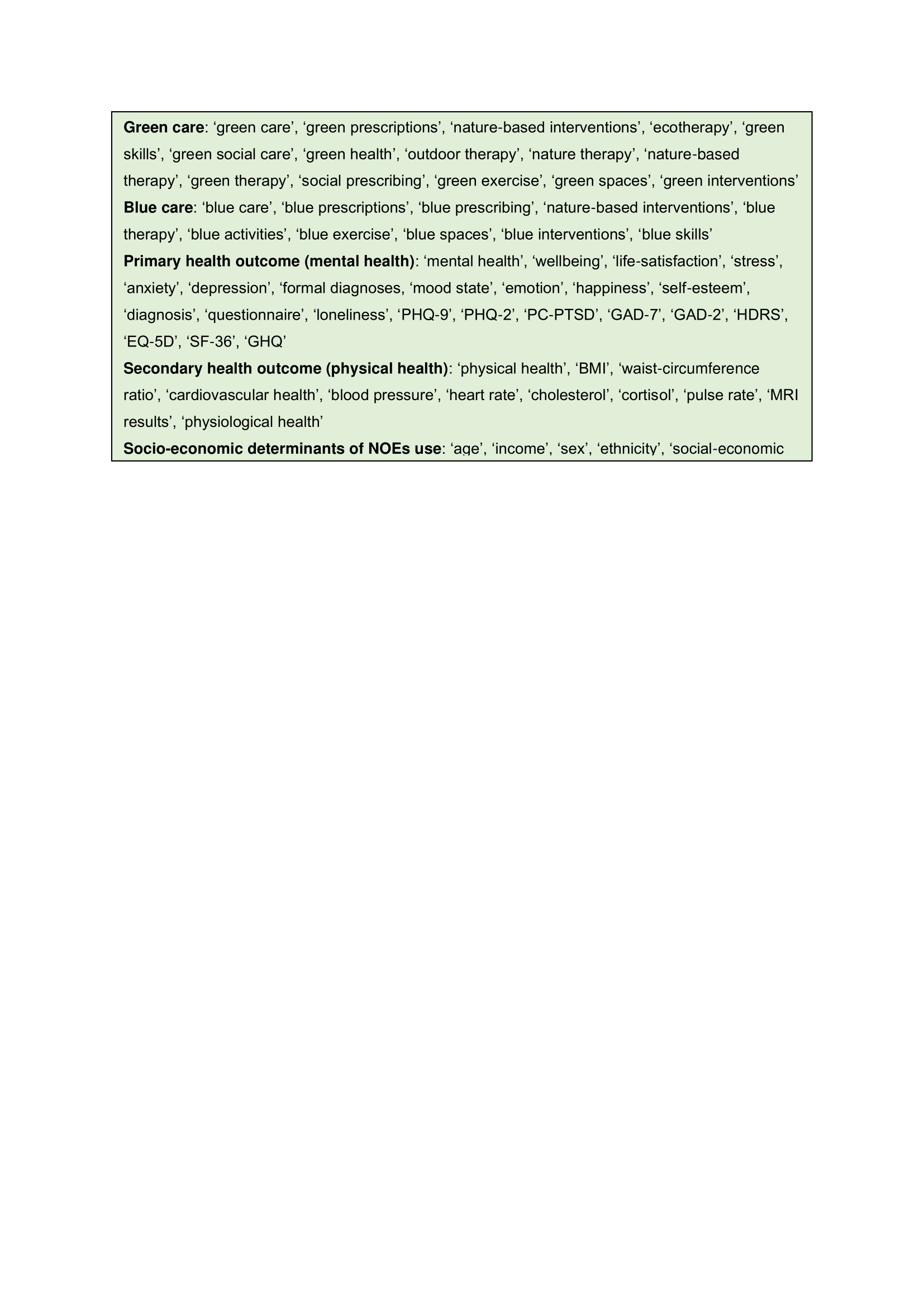

### Supplementary Figure 2

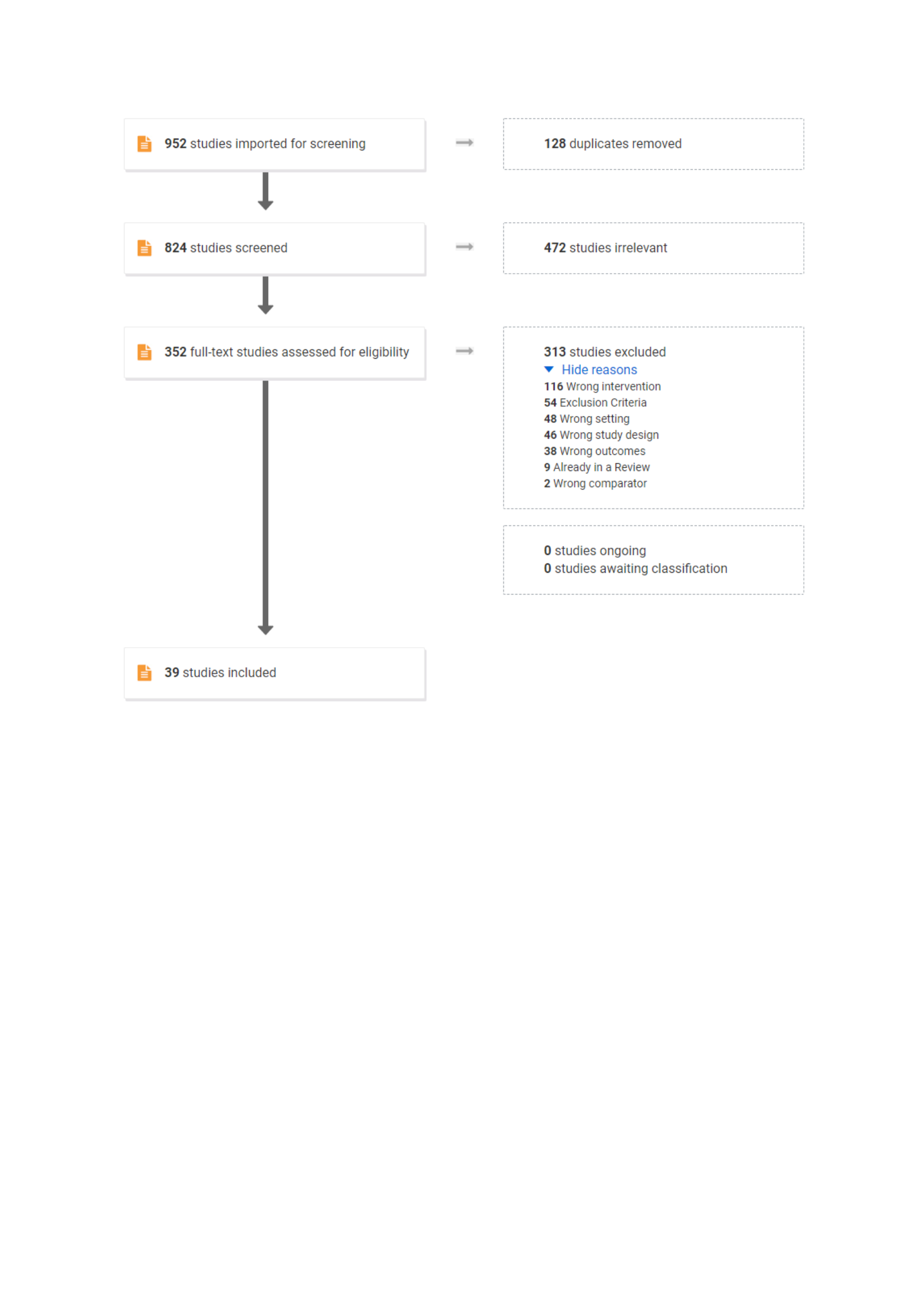

### Supplementary Figure 3

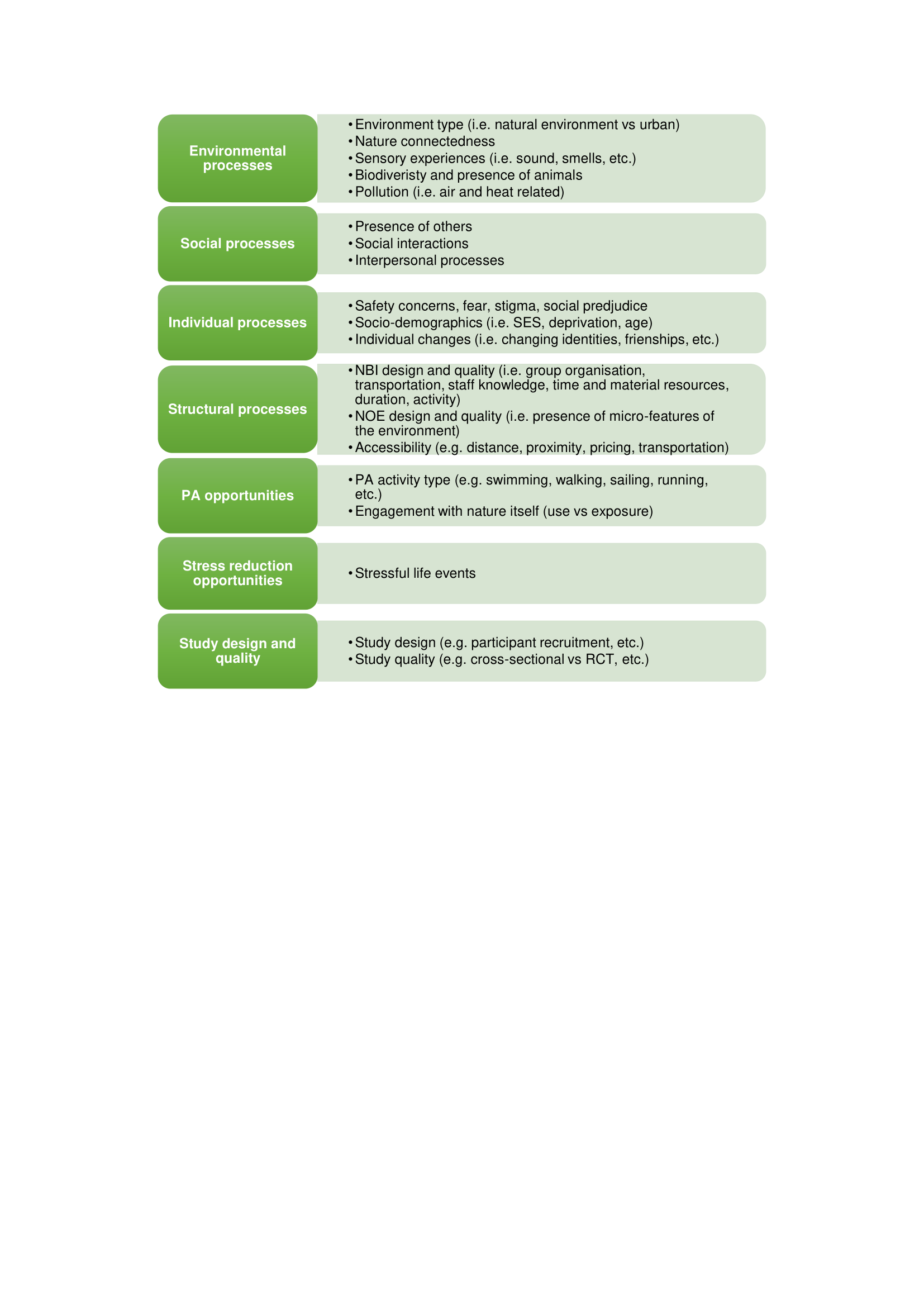

### Supplementary Table 1

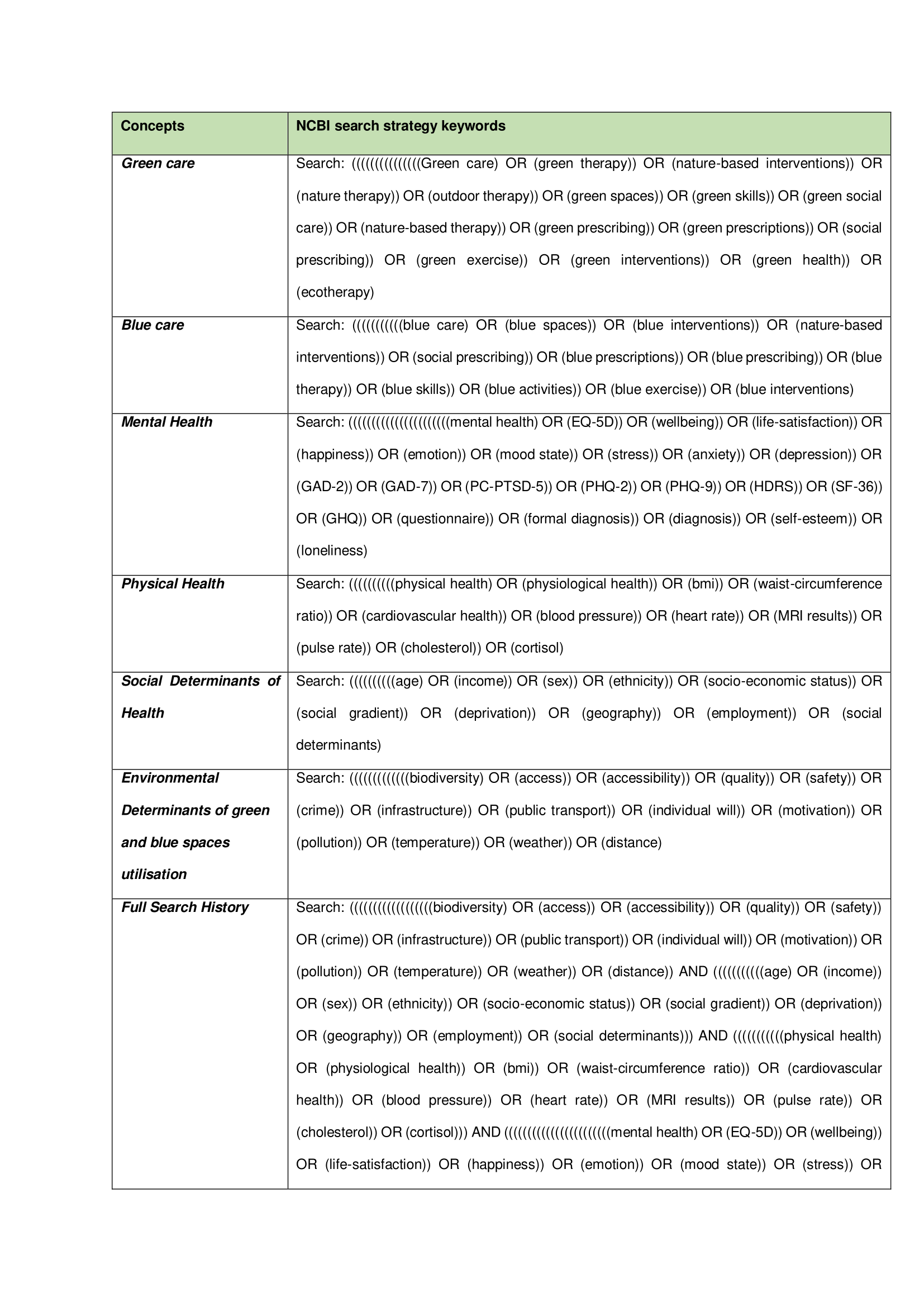

### Supplementary Table 2

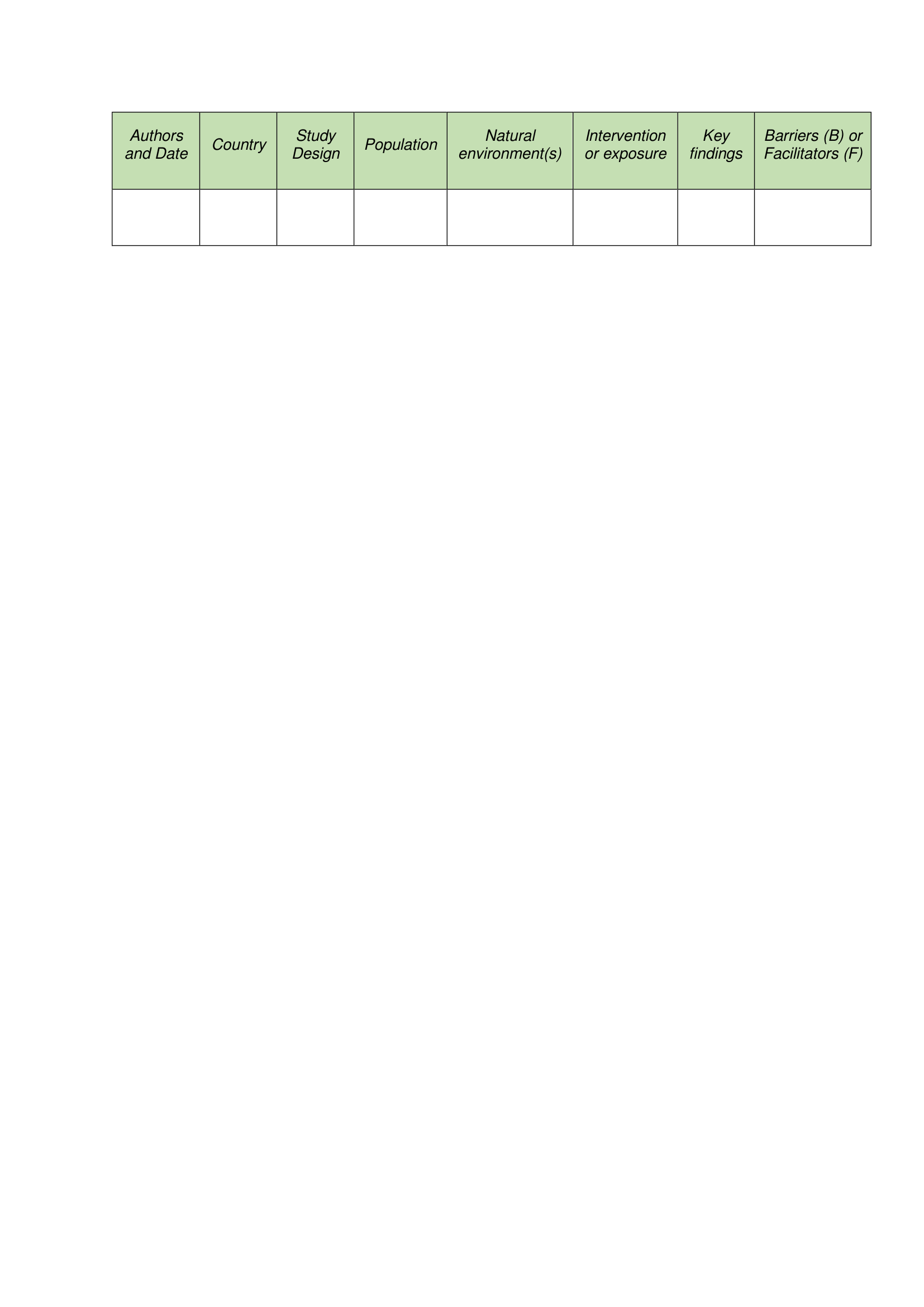
