## Supplementary Table 3 for "Enabling Health Outcomes of Nature-based Interventions: A Systematic Scoping Review"

| # | Authors and Date | Country | Study Design | Population | Natural environment(s) | Intervention or exposure | Key findings | Barriers (B) or Facilitators (F) |
|---|---|---|---|---|---|---|---|---|
| 1 | *Sprague N., Berrigan D., and Ekenga C. (2020)* | USA | Interventional study (non-randomized experiment) – with mixed-methods | **Children** (n=122; age range: 10-15-year-old) | Green spaces – urban forest parks, camping trips, urban farms, cave trips | NBE - Nature-Based Education | - Statistically significant positive changes in **STEM** capacity (+44%) and **HRQoL** (+46%) for participating students.<br>- Qualitative data highlighted the intervention's **educational** and **health** benefits. | - **Age** (F) - the older the child, the more active they will be, and the more benefits they will have on **HRQoL**. No effect for STEM.<br>- **Duration** - the longer tended to have more beneficial effects on **STEM capacity**.<br>- **Stressful life events** - the more one experiences, the less likely they will have benefits from NOEs. |
| 2 | *Arnberger A., Eder R., Allex B., Ebenberger M., Hutter H.P., Wallner P., Bauer N. , Zaller J. and Frank T.(2018)* | Austria & Switzerland | Interventional study (field experiment) – using mixed-methods | Adults (workers and university students) (n=22, age mean=26.7; SD=4.1) | Both - one urban city, two meadows (managed vs unmanaged) and river in mountain area | Wilderness expedition – walking and interacting (i.e. viewing) nature | - While differences measured on the physiological level between urban built and natural sites were **marginal** (on DBP not SBP), psychological measures showed **higher health benefits** of the **natural environments** | - **Environment type** (B/F) - river and alpine mountain meadow had the highest health benefits in terms of restoration, BP, and perception of beauty; but all sites recorded an improvement on calming and positive effect post-intervention. |

| | | | | | | | | |
|---|---|---|---|---|---|---|---|---|
| | | | | | | | compared to the built one. | |
| 3 | *Gargiulo I., Benages-Albert M., Garcia X. & Vall-Casas P. (2020)* | Spain | Observational study (exploratory fieldwork) – using qualitative methods | Adults stream users (N=30; F=14 and M=16, age range: 27-65+ years old) | Green – urban stream corridor | Leisure-Time Physical Activity (LTPA) | - Social and physical factors of the environment are perceived as either barriers or facilitators, with different nuances and importance, depending **on each type of user**.<br>- Also, for the same type of user, factors perception also depends on gender; whereby safety was important for women engaging in LTPA. | - **Safety** (B) – women reported lower level of use of blue/green spaces during LTPA if safety was a concern.<br>- **Environmental design** (B/F) - itineraries with **enhanced visibility,** higher **attendance**, pruning of dense vegetation and provisioning of **assistance** in case of need, all promoted engagement in LTPA.<br>- **Accessibility** (F) - stream accessibility and proximity to environments is conductive for LTPA.<br>- **Presence of others** (B/F) - having someone to share the experience with facilitated engagement. But |

| | | | | | | | | |
|---|---|---|---|---|---|---|---|---|
| | | | | | | | | can be a barrier when perceived as **safety risk** (i.e. walkers vs runners/cyclists and vice-versa).<br>- **Environment type** (F) |
| 4 | *Denton H. and Aranda K. (2019)* | UK | Observational study (ethnography) – using qualitative methods | Adults (regular swimmers and existing club members) (n=6; F=3 and M=6, age range: 38-73 years old) | Blue space - sea swimming club (Brighton, UK) | Sea swimming | - The swimmers found sea swimming **transformative**, (resulting in changes in the swimmer's experience of themselves); **connecting (**experiencing a sense of connection to nature, place and others); and **re-orientating** (as swimmers seemed to use this disruption to reconnect to what they consider is important), through **the disruption to the sense of time, space and body**, swimmers find alternative and | - **Physical activity** - engaging actively in the sea by swimming was critical to gaining health benefits (i.e. emotional and physical health)<br>- **Fear and stigma** - negative body image can impact one's engagement in sea swimming; but also fear of the challenges from the sea. |

| | | | | | | | | |
|---|---|---|---|---|---|---|---|---|
| | | | | | | | expanded perspectives about themselves and their world. | |
| 5 | *Finlay J., Franke T., McKay H. and Sims-Gould J. (2015)* | Canada | Observational study (participant observation) – using qualitative methods | Older adults (community dwellers; T1: N=27; T2: N=19, age range: 65-86 years old) | Both - urban parks in neighbourhood, with green and blue features | Walking | - Older adults have distinct **therapeutic relationships** with landscapes.<br>- Nature **can promote the physical, mental, and social health** of older adults.<br>- Blue space in particular embodies important therapeutic qualities for older adults. | - **Safety** (B/F)- can be experienced differently by people.<br>- **Accessibility** (B/F) - the least accessible, the worst the experience<br>- **Personal perception** (B/F) - the same place could evoke feelings from enjoyment to indifference to concern due to traffic, park maintenance, walkability, etc. |

| | | | | | | | | |
|---|---|---|---|---|---|---|---|---|
| 6 | *McEwan K., Richardson M., Sheffield D., Ferguson F.J. and Brindley P. (2019)* | UK | Interventional study (RCT) – quantitative methods | Adults (18+) residing in<br><br>Sheffield and owning a smartphone<br><br>(N=148) | Greenspace – urban park vs control (urban built) | Social prescription app on smartphone design to make people **notice** nature (while being in nature) | - Using a social prescription using a Smartphone app (noticing nature) resulted in statistically significant **improvements** in **wellbeing** for adults in general, and **clinically significant** improvements in wellbeing for those classed as having a mental health difficulty.<br>- These improvements were more pronounced in the **green space** condition, despite improvements still in control. | - **Environment type** (F) – both built and green environment yielded short-term benefits on wellbeing through **nature connectedness**; but only green space condition had sustained effects after one-month follow-up.<br>- **Previous experience with nature** (F) – from childhood or in the last year, both have positive effects on wellbeing.<br>- **Positive affect** (F) – predictor of wellbeing in green condition. |
| 7 | *Nicolosi V., Wilson J., Yoshino A.& Viren P. (2020)* | USA | Interventional study (field experiment) – using quantitative methods | Adults (undergraduates)<br><br>(n=63; F=31 and M=32; | Blue spaces – the coast vs control (urban side-road) | Coastal and urban walk | - Significantly higher average **perceived restoration** scores were associated with the natural (coastal) walk.<br>- **Coastal exposure, sound quality** | - **Perceived sound level** (F) - higher perceived sound level was a significant predictor of a restorative experience.<br>- **Environment type** (F) – if in natural environment, then increased |

| | | | | | | | | |
|---|---|---|---|---|---|---|---|---|
| | | | | age mean=20.4) | | | **and type** were rated as very good and more natural than the sidewalk respectively and were significant predictors of **a restorative experience.** | perceived restoration. |
| 8 | *Cheesbrough A.E., Garvin T., Nykiforuk C.I.J. (2019)* | Canada | Observational study (case study)– using qualitative methods | Adults and older adults (residents around one of the five selected NAP); (n=33; F=18 and M=15; age range=29-87 years) | Both – within five Natural Area Parks (blue and green features present) | Nature photography and reflection in nature | - **Proximity** to natural areas facilitated frequent and spontaneous visits.<br>- **Repeat visits** fostered intimacy with the space over time.<br>- Participants felt 'away from the city' while in the middle of the city.<br>- Participants reported **physical, spiritual and psychological therapeutic impacts**.<br>- Natural areas facilitated **connections to nature**, **self**, **companions**, and **others**. | - **Proximity** (F) - increase engagement **in physical activity** and therefore promotes improved health benefits.<br>- **Topography** - more difficult terrain were **motivational** for users of NAPs.<br>- **Sensory qualities** (F) - facilitated a **positive experience** of being in nature, and therefore allowed for this visit to be a **restorative experience**.<br>- **Safety** (B) - main barrier to using these areas (whether for fear of humans or wildlife). |

| | | | | | | | | |
|---|---|---|---|---|---|---|---|---|
| 9 | *Barton J., Bragg R., Pretty J., Roberts J., and Wood C. (2016)* | South Africa & Scotland | Interventional study (field experiment) – using quantitative methods | Adolescents (n=130; F=74 and M=57; age range: 11-18 years old) | Both – in a game reserve and a loch | Wilderness expedition in natural environments | - Environment, gender, and the length and location of expeditions significantly contributed to PPTs' changes in self-esteem (SE) and nature connectedness (NC).<br>- PPTs living in urban environments and going to local wilderness for a short duration will receive the same amount of benefits SE and NC as PPTs who live in a **rural location** and are immersed in a remote wilderness for longer. | - **Gender** (F) - males had higher self-esteem at start, but significant increase in SE for females at the end.<br>- **Duration** (F)- even short durations of expeditions can have benefits on nature connectedness and self-esteem. |

| | | | | | | | | |
|---|---|---|---|---|---|---|---|---|
| 10 | *Lanki T., Siponen T., Ojala A., Korpela K., Pennanen A., Tiittanen P., Tsunetsugu Y., Kagawa T. and Tyrväinen L. (2017)* | Finland | Interventional study (field experiment) – using quantitative methods | Adults (female volunteers<br><br>in Helsinki) (n=36;<br><br>age range: 30-60 years old) | Green space (vs control) - an urban forest, an urban park, and a built-up city centre | Each visit: 15 min of sedentary viewing; and 30min of walking | - Beneficial changes in **cardiovascular physiology** were observed in green environments.<br>- Specifically, **lower blood pressure** (viewing period only), **lower heart rate**, and **higher indices of heart rate variability**.<br>- **Large** urban park and **extensively managed** urban woodland had positive influence, but the overall perceived restorativeness was higher in the **woodland**.<br>- This may be explained by **stress relief** and **lower air pollution** and **noise exposure**. | - **Stress relief** (F) – the more relaxed one is in NOE, the better their health outcomes.<br>- **Air pollution** (B) – the higher the air pollution, the worst the health outcomes.<br>- **Noise exposure** (B) – higher noise engenders higher stress, and therefore lower health benefits. |

| | | | | | | | | |
|---|---|---|---|---|---|---|---|---|
| 11 | *Marselle M.R., Warber S.L. and Irvine K.N. (2019)* | UK | Observational study – using quantitative methods | Adults (volunteers) (N=1,516; age range: 55 years or older) | Both - natural environment (i.e., natural and semi-natural places, farmland, green corridor, coastal area, urban green space, or any mixture of the above) | Nature group walks | - Neither nature group walking, nor doing this frequently, moderated the effects of stressful life events on mental health.<br>- The positive associations of group walks in nature were at a greater magnitude than the negative associations of stressful life events on depression, positive affect, and mental well-being, suggesting an '**undoing' effect of nature group walks**. | - **Stressful life events (B)** – walking can help un-do stress associated with stressful life events by reducing depression and increasing positive affect and wellbeing.<br>- **Presence of others** (B) - can dampen buffering effect of nature on mental health.<br>- **Physical activity** (F) – mechanism by which individuals gain benefits from nature. |
| 12 | *PÁLSDÓTTIR A.M., STIGMAR K., NORRVING B., PETERSSON I.F., ÅSTRÖM M. and PESSAH-RASMUSSEN H. (2020)* | Sweden | Interventional study (RCT) – using quantitative methods | Adults and older adults (stroke survivors) (n=101; F= | Green space - Alnarp Rehabilitation Garden (Nature Area (informal and non-cultivated) and the Cultivation and Gardening | Nature-based rehabilitation (NBR) using horticultural therapy | - The patients with sub-acute stroke were highly **compliant** with the intervention. The participants in both the intervention and control groups **improved**. | - **Weather** (B) - not suitable NBR in bad weather.<br>- **Access** (B) - acted as barrier to participation in NBR for some PPTs due to longer travel time to the garden. |

| | | | | | | | | |
|---|---|---|---|---|---|---|---|---|
| | | | | 61 and M=40; age range: 50–80 years old). | Area (formal land cultivated)) | | - However, **no statistically significant differences** in improvement were found between the intervention and control groups for any of the outcome measures.<br>- **Fatigue decreased** to a value below the suggested cut-off for mental fatigue (< 10.5) in the intervention group, but not in the control group. | |
| 13 | *Pratiwi P.I., Xiang Q. and Furuya K. (2019)* | Japan | Interventional study (field experiment) – using quantitative methods | Adults and older adults (local residents) (n=12 in spring; F=6 and M=6; mean age: 66.4) and (n=12 in summer; F=7 | Green spaces – across three sites: urban city site and two viewing spots in urban park | Viewing cherry blossom trees and fresh greenery in urban parks VS urban city in Spring and August | - **Viewing cherry blossoms and fresh greenery** in urban parks led **to lower blood pressure** in spring and early summer than viewing city areas in spring and early summer.<br>- The results of this study | - **Seasons** (F) **-** positive mood states were higher in spring as well as lowered mood disturbances; whereas state-anxiety levels were lower in early summer.<br>- **Environment type** (F) - green spaces > urban city for physiological and |

| | | | | | | | | |
|---|---|---|---|---|---|---|---|---|
| | | | | and M=5; mean age: 65.75) | | | suggest that viewing urban parks results in physiological and psychological relaxation. | psychological effects of NBI.<br>- **Biodiversity and surrounding features** (B/F) (i.e. mosquitoes, sun, temperature, etc.) - thought to have increased heart rate and modified BP measurements, when viewing urban parks in both seasons.<br>- **Presence of water** (F) - associated with a significant **positive effect** and high perceived restorativeness.<br>- **Traffic** (B) (i.e. people or noises from vehicles) - could be responsible for altering BP measures in urban city. |

| | | | | | | | | |
|---|---|---|---|---|---|---|---|---|
| 14 | *Byström K., Grahn P. and Hägerhäll C. (2019)* | Sweden | Interventional study (field experiment) – using qualitative methods | Children (with disabilities, i.e. autism)<br>(n=9; mental age range: 4–6 years) | Both – immersion in a farm and surrounding nature (no specifications) | **KOMSI treatment** – nature therapy for treating children with disabilities (i.e. horseback riding, free play, etc.) | - The intervention led researchers to conclude on three key benefits of the intervention: 1) reduce stress and instill calm, 2) arouse curiosity and interest, and 3) attract attention spontaneously.<br>- These three perceived benefits are related to **vitality forms**. It is argued that the **vitality forms** from nature and animals are favorable for effecting development-promoting interactions with a therapist. | - **Therapeutic environment (F/B)** - if in nature it can trigger positive or negative responses for the child (i.e. not all autistic children would appreciate being out)<br>- **Presence of animals** (F) - for this subgroup, animals and nature facilitated communication and alleviated stress. |
| 15 | *Ana BY, Wanga D., Liua XJ., Guanb HM., Wei HX. and Renb ZB. (2019)* | China | Interventional study (field experiment) – using quantitative methods | Adults (undergraduates in horticulture) | Green spaces – across three types of forests: 1) Maple,<br>2) Birch<br>3) Oak | Forest Bathing in three types of forests | - This study looked at the relationship between environmental factors (temperature, RH, light intensity, and light spectrum) | - **Tree species** (F) - maple> oak> birch for HR improvements. Yet, birch forests still had HR improvements, and was the only one to demonstrate that at |

| | | | | | | | | |
|---|---|---|---|---|---|---|---|---|
| | | | | (n=13; M=7 and F=6; mean age: 21 years old) | | | and physiological changes (SP, DP, and HR).<br>- Pre--forest-bathing **temperature** and **spectrum** can impact the response of blood pressure due to the "law of the initial value".<br>- HR was influenced positively by visits to maple >oak>birch trees.<br>- Authors recommend visitors to walk in **maple** forests to obtain cardiovascular and autonomic nervous system well-being. | lower levels of BP to begin with.<br>- **Temperature** (B) - Pre-forest bathing temperature can negatively impact the response of BP if PPTs felt too cool and moist.<br>- **Light spectrum** (B) - pre-forest bathing spectrum can negatively impact the response of BP if high G/B ratio are too extreme. |
| 16 | *Leavell M.A., Leiferman J. A., Gascon M., Braddick F., Gonzalez J. C. and Litt J. S. (2019)* | USA | Literature review | Across age groups | Both – across several types of natural environments | Several types of NBIs being reviewed here - i.e. water rafting, horticulture, green exercise | - Nature-based social prescription increases **social connectedness** and influences **physical health** and **mental well-being** by certain | - **Intrapersonal processes** (F) - give way to social connections and longer-term health outcomes.<br>- **Interpersonal processes** (F) - improves social connections and |

| | | | | | | | | |
|---|---|---|---|---|---|---|---|---|
| | | | | | | | intrapersonal, interpersonal, and environmental pathways.<br>- NBI practice represents a low cost, creative intervention to **strengthen social networks, reduce stress, and facilitate social connectedness** among participants and providers. | health outcomes by promoting social involvement, relatedness, and shared learning.<br>- **Environmental processes** (B/F) – such as access to nature, perceived neighbourhood attachment, and perceived aesthetics. |
| 17 | *Hunter R.F., Cleland C., Cleary A., Droomers M., Wheeler B.W., Sinnette D., Nieuwenhuijsen M.J. and Braubachi M. (2019)* | USA, Australia, UK | Meta-narrative<br>evidence synthesis | Across age groups | Urban green spaces – i.e. urban parks, rooftops, parking lots, etc. | Any NBI intervention that has only physical changes to the UGS or with health promotion to tackle inequalities | - There was strong evidence for: 1) **park-based and greenway/ trail interventions** employing a **dual** approach (i.e. a physical change to the UGS and promotion/ marketing programmes);<br>2) **Greening of vacant lots** which reduced stress and social benefits (e.g. | - **Changes to the built environment in parks** (with dual-approach) (F) - provision of signage and community garden, improvements in existing playing fields, replacement of old playground equipment, installation of outdoor gyms, improved footpaths and clearing of rubbish and vandalism all increased |

| | | | | | | | | |
|---|---|---|---|---|---|---|---|---|
| | | | | | | | reduction in crime, increased perceptions of safety); 3) **Greening of urban streets** and SuDS for managing storm water had environmental benefits as well. | individual's park use, physical activity and the latter two improved QoL and perception of safety.<br>- **Proximity** to newly developed walking/ cycling routes (F) - increased use of these UGS.<br>- **Greening of vacant lots** (F) – reduced perception of unsafe environment and bolster use of these UGS. |
| 18 | *van den Bosch M. and Sang O. (2017)* | Not provided | Systematic review (of reviews) | Across age groups | Both - (i.e. green infrastructure, biodiversity, blue environments, etc.) | Interventions in urban natural environments | - There is strong evidence on the effect of urban nature **on affect state**.<br>- There is strong evidence on the effect of urban nature on **urban heat reduction**.<br>- **Positive affect and heat reduction** can mediate urban nature's effect on **mortality**. | - **Micro-features** (F)<br>- **Conditions of natural environments** (B/F)<br>- **Perceived quality** (F)<br>- **Accessibility** (B/F)<br>- **Safety** (B) |

| | | | | | | | | |
|---|---|---|---|---|---|---|---|---|
| 19 | *Houlden V., Weich S., de Albuquerque J.P., Jarvis S. and Rees K (2018)* | Europe USA Canada, Australia | Systematic review | Adolescents, adults, and older adults | Green spaces – mixed definition that encompasses vegetated areas and/or wilderness | Not interventions per se, but includes studies with walking in GS as measure for visits to GS | - There was **adequate evidence** for associations between the **amount of local-area greenspace and life satisfaction** (hedonic wellbeing), but **not personal flourishing** (eudaimonic wellbeing).<br>- Evidence for associations between **mental wellbeing and visits to greenspace, accessibility, and types of greenspace** was limited.<br>- There **was inadequate evidence** for associations with views of greenspace and connectedness to nature. | - **Views of greenspace** (B/F) **-** if looking at unpleasant urban/rural views it will have negative association with mental health.<br>- **Connection with nature** (F) - the more connected one is with nature, the more health benefits (i.e. life satisfaction, happiness, affect, QoL) one will experience. This is modulated by **being actively engage** in nature, however.<br>- **Visits to greenspaces** (F) - active immersion in wilderness was found to lead to greater happiness, affect and attention. |

| | | | | | | | | |
|---|---|---|---|---|---|---|---|---|
| 20 | *McCormick R. (2017)* | USA, Spain and others not specified | Systematic review | Children (age range: 0-18) | Green spaces – wooded playgrounds, natural habitats, gardens, etc. | Not interventions per se but does include studies who used walking in NOE as measure for visits to green space. | - **Access** to green space is important to the **mental well-being**, **overall health,** and **cognitive development** of children. It promotes **attention restoration**, moderates the impacts of **stress**, improves **behaviours** and **symptoms of ADHD** and was even associated with **higher standardized test scores**. | - **Proximity to GS** (F) - only passive exposure but the closer one lives to nature, the better health outcomes they have.<br>- **Physical activity** in NOE (F) - **walking** in nature for children vs in urban environment led to improved attention and spatial working memory - which can help children with ADHD focus better. |
| 21 | *Barrett. J., Evans S. and Mapes N. (2019)* | UK | Literature review | Older adults (residents of dementia care settings) | Green space – garden areas within dementia care settings | Green dementia care - includes many type of NBIs (i.e. horticulture, walking, gardening, etc.) | - Compelling evidence for **several health and wellbeing benefits** associated with green dementia care (i.e. improved wellbeing, social interactions, stress-reduction and restorative effects, self- | - **Safety concerns** (i.e. fear of falling in garden) (B)<br>- **Staff attitudes** and **lack of staff education** and awareness (B)<br>- **Social prejudice and stigma** (B)<br>- **Limited staff** to accompany residents and **limited resources (B)**<br>- **Weather** (B) |

| | | | | | | | | |
|---|---|---|---|---|---|---|---|---|
| | | | | | | | worth and confidence.)<br>- Evidence base is stronger regarding the **barriers and facilitators** to accessing nature for this population - **staff education and care culture** is critical to the success and effective use of the garden for such residents.<br>- **Design** of the outdoor space need to ensure that these spaces are **visually and physically accessible** for its residents. | - **Self-perception of being too old** and **lack of confidence** (B)<br>- Poor physical and visual access<br>- **Poor garden design** (B) (i.e. lack of resting places and weather protection)<br>- **Care culture NOT person-centred** (B) |
| 22 | *Ottoni C.A., Sims-Gould J., Winters M., Heijnen M. and McKay H.A. (2016)* | Canada | Observational study (participant observation) – using | Older adults (60+) (n=28; F=17 and M=11, age range: 61-89, in 2012; n | Both areas – three areas in parks in Vancouver with features of GS and BS | Physical activity in nature - recorded as step counts/day (mean) | - **Neighbourhood environments** influence health and well-being as people age.<br>- There are strong interconnections between **built** | - **Amenities** (F) – i.e. benches, seen as a necessity to promote social interactions and positive experiences.<br>- **Ability to engage in other type of activities** (F) – i.e. |

| | | | | | | | | |
|---|---|---|---|---|---|---|---|---|
| | | | qualitative methods | =22, F=12 and M= 10; in 2014) | | | **and social environments**.<br>- **Microscale features** can enable older adults' to accommodate to their abilities.<br>- **Benches** can promote mobility and social connectedness for older adults.<br>- **Microscale features**, like benches, are a prudent investment for communities. | family or friends activities, going to the pub, going to the gym featured more prominently than benches in relation to their mobility.<br>- **Injury** (B) - to use the outdoor environments.<br>- **Wildlife** (F) - promoted feelings of enjoyment and calmness. Also helped in creating routines/familiarity with these spaces.<br>- **Presence of other people** (B/F) - a negative experience for older adults if too many people use benches. But seeing people around them also provided positive feelings opportunities for social interactions.<br>- **SES** (B) - **accessibility and availability** of GS and BS was more common for older adults with higher |

| | | | | | | | | |
|---|---|---|---|---|---|---|---|---|
| | | | | | | | | social and economic status. |
| 23 | *Howarth M., Rogers M., Withnell N. and McQuarrie C. (2018)* | UK | Observational study (cross-sectional) – using mixed-methods | Adults and older adults (suffering from mental disorders (n=47; age range: 35-68 years and average age: 53.2 years) | Green space – garden area created by social enterprise | Therapeutic horticulture | - Quantitative findings showed that participants were working towards **self-reliance**. Qualitative data found similar results.<br>- **Mental health recovery programme** enabled participant **integration** into the community through providing a space to **grow and build self-confidence** while reengaging with society.<br>- The results suggest that using therapeutic horticulture as | - **Positive staff attitudes** (F) - welcoming and non-judgmental attitudes promoted wellbeing and social connection for this population. It also helped people feel safer.<br>- **Activities as a new purpose** (F) – engaging in nature itself improved wellbeing and allowed people to feel a sense of purpose. By developing new skills people felt more confident in their own self and their employability.<br>- **Presence of others** (F) - improved sense of purpose and recovery, as everybody shared |

| | | | | | | | | |
|---|---|---|---|---|---|---|---|---|
| | | | | | | | an intervention within the mental health recovery programme can **support people with mental health problems** to re-engage socially. | same/similar experiences; this helped them move beyond their diagnosis. It also provided opportunities to re-engage with society. |
| 24 | *Kabisch N., Matilda van den Bosch M. and Lafortezzae R. (2017)* | U.S., Germany France Spain Denmark, Bulgaria, Austria, Sweden, UK, Japan, Canada and China | Systematic Review | Children and the elderly | Both – features of both GS and BS | Some studies included interventions / active engagement with nature | - There is a tendency for a positive association between urban green and blue spaces and reduced risk factors related to urbanization for children and the elderly as well as the promotion of health-related behaviours and subsequent positive health outcomes.<br>- But the evidence is weak and the results are somewhat inconsistent. | - **Socioeconomic factors** (B) (i.e. deprivation, income, educational level, unemployment) - the lower one's household, the worst their health outcomes, and the lower the relationship between health and nature.<br>- **Air pollution** (B) - act as mediator of the relationship between nature and health - but not if elderly engage actively in NOE (i.e. gardening).<br>- **Heat-related pollution** (B) - the higher the heat in parks, the less use and the worst health outcomes, |

| | | | | | | | | |
|---|---|---|---|---|---|---|---|---|
| | | | | | | | | specifically for the elderly.<br>- **Proximity/distance** (F/B) - proximity can modify effectiveness of NBIs. |
| 25 | *Shin J.C., Parab K.V., An R. and Grigsby-Toussaint D.S. (2020)* | USA, Australia, Canada, Spain, UK, Netherlands, Lithuania | Systematic review | Across age groups | Green spaces – neighbourhood greenness, visits to GS, engagement in activities related to GS | Several interventions included: walking, gardening, work environment | - **Green space exposure** (through active engagement) is associated with **better sleep quality and quantity.**<br>- Authors suggest **green exercise** and **therapeutic gardening** for future interventions. | - **Time of day for activity** (F) – afternoon walking > morning walks.<br>- **Type of environment (F)** - outdoor > indoor interventions on sleep latency.<br>- **Behavioural preferences** (F) - people had better sleep on weekdays with lower exposure to green spaces. Also preferred vaster expenses of greenspace on weekends and for longer than during weekdays. |
| 26 | *Lakhani A., Norwood M., Watling D.P., Zeeman H. and Kendall E. (2019)* | USA, Norway, Netherlands, Australia, Korea, Japan | Systematic review | Adults and older adults (suffering from neurological disability: | Both – includes studies with features from both green and blue environments | Several interventions included: gardening, green care farming, wilderness therapy, | - Given the limited research to date, and the diversity of nature specific activities, it is **not possible to establish** | - **Environment type** (F) - for care farming, evidence is mixed on social health.<br>- **Garden design** (F) - gardens often well arranged, walled |

| | | | | | | | | |
|---|---|---|---|---|---|---|---|---|
| | | | | dementia, stroke,<br>and brain injury) | | forest therapy | **definitive conclusions** around the efficacy of engaging with nature specific activities on the **psychosocial health** of people with neurological disability.<br>- At best, findings clarify that **engaging** with natural environments contribute to favourable **emotional health outcomes and social health outcomes** for people with dementia. | and preferably in connection with a shielded dementia care unit, may also improve agitation among people with dementia.<br>- **Mobility** (B) **-** impact of wander gardens on agitation reduction was lower if PPT had ambulatory issues.<br>- **Presence of caregiver** (F) **-** brought positive emotional health outcomes in patients with dementia, when entering garden.<br>- **Short-term plants** (F) –associated with improved social health vs long-term plants. Possibly due to faster harvesting capacity.<br>- **Active engagement** (F) **-** psychological health has been favourably impacted only when activities in nature |

| | | | | | | | | |
|---|---|---|---|---|---|---|---|---|
| | | | | | | | | involve active engagement. |
| 27 | *Kondo M.C., Fluehr J.M., McKeon T. and Charles C. Branas C.C. (2018)* | USA, UK, Netherlands, Canada, Lithuania, Denmark, Germany, Finland, Japan, Italy, Spain | Systematic review | Across age groups | Green spaces – natural environment | Several types of interventions including: viewing nature, walking, exercising, gardening | - This review of experimental, quasi-experimental, and longitudinal studies found **evidence of a positive association** between urban green space and **attention**, **mood**, and **physical activity**, and **negative association with mortality, short-term cardiovascular markers (heart rate), and violence**.<br>- In most cases, it is not possible to observe patterns of findings of association between urban green space exposure and health outcomes (i.e. birth outcomes, | - **Environment type** (F) - natural environment > urban built environment for attention, general health, cardiovascular outcomes (i.e. HR, HRV), mood and emotions (i.e. specifically urban woodlands for restoration).<br>- Biodiversity (F) - found to improve **mood** and **emotions** but is mediated by length of park visit and perceived restoration.<br>- Physical activity (F) - engagement in PA in nature was positively associated with health outcomes in experimental studies VS observational studies. |

| | | | | | | | | |
|---|---|---|---|---|---|---|---|---|
| | | | | | | | stress, BP, cancer, diabetes, etc.). | |
| 28 | *Callaghan A., McCombe G., Harrold A., McMeel C., Mills G., Moore-Cherryb N. and Cullen W. (2020)* | Australia, USA, UK, Bulgaria, Belgium, Denmark, Netherlands, Serbia | Scoping review | Across age groups | Green spaces – urban parks, neighbourhood greenness | Several interventions included: horticultural therapy, walking, viewing from indoors | - The majority of studies found a **positive association** between GS and mental health.<br>- Policies to increase urban green space may have **sustainable public health benefits**. | - **Ethnicity** (B) - South Asian children living in more deprived areas and with lower access and quality of GS had more behavioural difficulties VS white British children.<br>- **Deprivation** (B) - quality and access to greenspaces is lower in deprived and lower-income communities, |

| | | | | | | | | |
|---|---|---|---|---|---|---|---|---|
| | | | | | | | | leading to worse health outcomes. |
| 29 | *Koselka E.P.D., Weidner L.C., Minasov A., Berman M.G., Leonard W.R., Santoso M.V., de Brito J.N., Pope Z.C., Pereira M.A. and Horton T.H. (2019)* | USA | Interventional study (pilot study) – using quantitative methods | Adults (undergraduates, graduates and employees) (n=37; 18–35 years; age mean=22.9 and SD=4.6) | Green | Walking in nature across 3 settings: forest; along roadside, activities of daily living | - This study has found that **moderate-intensity walking** in a forested environment had a **positive impact on psychological health**.<br>- This suggests that completing **physical activity** in greenspaces **amplifies beneficial acute psychological responses** and yields greater improvements in **mental health** than does activity completed indoors or in a built urban environment. | - **Type of environment** (F) - forest walking >roadside> daily activities for positive/negative affect, perceived stress, and anxiety.<br>- **Physical activity** (F) - walking in general brought improved mental health but amplified in forest environment. |
| 30 | *Zufferey J. (2016)* | Japan, Australia, China, USA, New | | | Both – GS and BS elements (not specific) | Not specified per se, but includes studies with walking/ | - This literature review shows moderate to strong empirical evidence for the | - **Age** (F) - children and young adults seemed to benefit more from exposure to GS and |

| | | | | | | | | |
|---|---|---|---|---|---|---|---|---|
| | | Zealand, Canada | Systematic review | Across age groups | | exercise in GS and BS | positive influence of **contact** with green and blue spaces and **mental and physical health** and **low evidence** for influences on **social cohesion**.<br>- It also shows that health impacts may vary according to the **population group** considered (e.g. children, people with low socio-economic status who benefit more from exposure to these environments). | BS, especially via physical activity; which had combined effects on physical and mental health.<br>- **SES** (B/F) - lower SES households tended to have more health benefits associated with exposure to green and blue spaces.<br>- **Type of environment** (F) - natural environments > urban built on emotional wellbeing. |
| 31 | *Costello L., McDermott M-L., Patela P. and Dare J. (2019)* | Australia | | | Blue – the ocean and surrounding beaches | Ocean swimming | - All the ocean swimming groups studied were united by their **routine** of beach swimming, by their **love of the ocean**, and their conviction that | - **Type of environment** (F) - swimming in the ocean VS public/private pool.<br>- **Biodiversity** (B/F) - when seeing fishes, dolphins, whales, etc. people experienced |

| | | | | | | | | |
|---|---|---|---|---|---|---|---|---|
| | | | Observational study (ethnography) – using qualitative methods | Older adults (self-organised ocean swimmers) (n=10; F=7 and M=10; age range: 55-80+ years) | | | their ocean swimming practice **as part of a group** was beneficial for their social connectedness, wellbeing and physical and mental health. | positive emotions and pleasurable experiences. However fear of sharks led to negative emotions, despite increasing social cohesion.<br>- **Type of activity** (F) - swimming > other type of outdoor exercise, as it was low-impact. It would also help in alleviating stress.<br>- **Weather** (F) - despite cold and rainy weather, PPTs would still engage in swimming, as their commitment to the group was the priority.<br>- **Group membership** (F) - PPTs recognised that they would not derive the same enjoyment, pleasure and health benefits w/o group. |
| 32 | *Birch J., Rishbeth C. and Payne S.R. (2020)* | UK | | Adolescents and adults (n=24; | Both – urban parks in Sheffield (UK) | Arts workshop and interviews in nature | - Deteriorating landscapes, young people's shifting identities and perceived time pressures | - **Poor quality of urban environment** (B) – or urban deprivation, was more impotant than |

| | | | | | | | | |
|---|---|---|---|---|---|---|---|---|
| | | | Observational study (case study) – using qualitative methods | F=14 and M=10; age range: 17-27 years, with n=9 experiencing mental difficulties and n=15 living in an area of urban deprivation). | | | disrupted support.<br>- Overall young people expressed how urban nature encounters were experienced **as accepting and relational**, offering a **stronger sense of self**; feelings of escape connection and care with the human and non-human world. | ethnicity and SES across PPTs.<br>- **Presence of others** (F/B) - having someone with you during a visit to an urban environment was positively experienced (i.e. wanting someone to share experience with), or negatively experienced (i.e. wanting to be alone).<br>- **Individual factors** (B/F) - pressures, changing priorities and changing friendships all have their mediating role. |
| 33 | *Wood E., Harsant A., Dallimer M., ronin de Chavez A,, McEachan R.R.C. and Christopher Hassall (2018)* | UK | Observational study (cross-sectional) – using quantitative methods | Adults and older adults (users of local parks in deprived areas) (n=128; F=59 and M=69; age range: 18-76+ years) | Green | Visits to greenspaces -<br>survey conducted at the<br>park entrance | - Authors found that **biodiversity** and site facilities were positively correlated within urban parks. However, we found that only **biodiversity was related to perceptions of psychological restoration** amongst a multi- | - **Biodiversity** (F) - the more biodiversity a park had; the more people would benefit from psychological restoration.<br>- **Amenities** - positively correlated with urban parks, but no effects on psychological restoration. |

| | | | | | | | | |
|---|---|---|---|---|---|---|---|---|
| | | | | | | | ethnic group of PPTs.<br>- These findings suggest that urban planners should aim to enhance **ecological diversity** in urban green spaces. | - **Ethnicity** - no effects found. |
| 34 | *Corazon S.S., Sidenius U., Poulsen D.V. , Gramkow M.C. and Stigsdotter U.K. (2019)* | Europe, Asia, Australia | Systematic Review (without meta-analysis) | Adults and older adults<br>. | Both – all types of outdoors natural green environments | All types of sedentary and light exercise activities, in all time durations in nature | - The synthesis of the results points towards outdoor, nature-based exposure having a positive effect on different **emotional parameters**, related to stress relief. The studies into physiological measures showed more equivocal results.<br>- The general use of **self-referred individuals** imposes a potential strong bias. | - **Type of environments** (F) – natural environments vs control had positive association with emotional outcomes (i.e. positive affect, perceived stress and wellbeing/QoL), and negative association with negative affect. This could not be found for physiological measures – (too heterogeneous). |

| | | | | | | | | |
|---|---|---|---|---|---|---|---|---|
| 35 | *Maund Pgreen.R., Irvine K.N., Reeves J., Strong E., Cromie R., Dallimer M. and Davies Z.G. (2019)* | UK | Interventional study (pilot study) – using mixed-methods | Adults and older adults (already registered with<br><br>the community mental wellbeing service and diagnosed with depression and/or anxiety) (n=16; F=8 and M=8) | Blue – wetlands | **Wetland NBI** – guided walking, bird watching or other activities (i.e. canoeing) done in nature over a six-week period. | - There were significant improvements in **mental health** across a range of indicators, including **mental wellbeing, anxiety, stress and emotional wellbeing**. Participants and healthcare professionals cited additional outcomes including **improved physical health** and **reduced social isolation**.<br>- The wetland site provided a **sense of escape** from participants' everyday environments, facilitating relaxation and **reductions in stress**.<br>- Wetland **staff knowledge** of the natural world, | - **NBI design** (F) - to be successful, NBIs need to take into account transportation, staff knowledge and group dynamics.<br>- **Biodiversity** (F) - the presence of water, diverse wildlife and the inherent peacefulness of wetlands were positively experienced with the intervention.<br>- **Session content** (B/F) - most PPTs preferred if there was only ONE activity vs many → less stress and anxiety that way. |

| | | | | | | | | |
|---|---|---|---|---|---|---|---|---|
| | | | | | | | **transportation** and **group organisation** also played a role in the intervention's success. | |
| 36 | *Benjamin-Neelon S.E., Platt A., Bacardi-Gascon M., Armstrong S., Neelon B., Jimenez-Cruz A. (2019)* | Mexico | Observational study (cross-sectional) – using quantitative methods | Children (n=102, age rage: 3-5 years; in Ensenada (M=29 and F=21) and in Tijuana (F=27 and M=25) | Green spaces – urban parks in two cities (Tijuana and Ensenada) | Time spent in GS (measured with GPS) | - **Greater time** in greenspace was associated with **decreased sedentary time**.<br>- **Greater time** in greenspace was associated with **increased physical activity**.<br>- Associations were mainly driven by children in **Tijuana** compared to Ensenada.<br>- Time spent in greenspace was **not associated** with body mass index **(BMI) z-score.** | - **Duration** (F) - the greater the time spent in greenspace, the less sedentary time these children will experience, but also the greater their MVPA will be (physical activity). |
| 37 | *Coventry P.A., Neale C. Dyke A., Pateman R.* | UK | | Adults and older adults (conservation volunteers) (n=45; F=20 | Green spaces – across three sites: Askam Bog, St Nicks natural reserve and a large green field with | Three interventions: group walking, conservation | - Undertaking **purposeful activity** in public green space has the potential to **promote health** and prevent | - **Location** (F)- effects differed for **stress** across locations, meaning that the location of the GS, over the type of activity, was |

| | | | | | | | | |
|---|---|---|---|---|---|---|---|---|
| | *and Cinderby S. (2019)* | | Interventional study (field experiment) – using mixed-methods | and M=25; age mean: 43.8) | surrounding woodland, adjacent to a semi-urban housing development | , citizen science | **mental ill health**. Undertaking such activities in locations where people have the **most connection** might confer additional benefits.<br>- **Social interaction, physical activity and restoration** were all implicated as potential mechanisms by which activities in public green spaces might lead to improved mental health. | an important factor in reducing stress - which was explained by an enhanced **place attachment** and **place identity** at this location.<br>- **Type of activity** (F) - although not shown quantitatively, **conservation and citizen science** were both associated with deeper **sense of purpose** by providing **learning opportunities**, and because it conferred **co-benefits** to health, wellbeing and to nature itself. |
| 38 | *Britton E., Kindermann G., Domegan C. and Carlin C. (2018)* | Europe, USA, Canada, New Zealand, Israel | Systematic review | Across all age groups – but with pre-existing condition | Blue space – wilderness, sea, urban/ semi-urban areas (beach, city), or mix of these | Several interventions included in BS: surfing, Dragon Boat Racing (DBR), sailing fly fishing kayaking, canoeing, at the beach, swimming, | - Blue care have **direct benefit** mental health and psycho-social wellbeing.<br>- There was also evidence for **greater social connectedness** during and after interventions, but results were **inconsistent** | - **Access** (F/B)<br>- **Lack of resources/ equipment** (B)<br>- **Fears/stigma** (B) - associated with personal abilities, level of fitness, environment, social and cultural norms, diagnosis of illnesses and level of appropriate training for those |

| | | | | | | | | |
|---|---|---|---|---|---|---|---|---|
| | | | | | | (as part of a kayaking intervention), and scuba diving | and mixed; with very few findings for physical health.<br>- Findings suggest how **activities** in BS, rather than particular **qualities** of BS, might contribute to rehabilitation and health promotion. | delivering intervention.<br>- Gender<br>- **Seasickness** - due to poor weather (B)<br>- **Fatigue/tiredness** - post-intervention (B) |
| 39 | *Saadi D., Schnell I., Tirosh E., Basagaña X. and Agay-Shay K. (2020)* | Israel | Interventional study (field experiment) – using quantitative methods | Adults (women specifically) (n=120; age range: 20-35, from two small cities in the north of Israel, of whom n=48 were Arab and n=24 were Jewish women (n= 72) | Green spaces – across Afula-urban park, Afula-city center, Afula-residential area, Nazareth urban park, Nazareth-city center and Nazareth-residential area VS home (control) | Viewing and waling in nature while sitting on benches across 6 different sites | - **Visits to urban parks** compared to staying in the home environment had beneficial short-term changes in **psychological**, **physiological**, and **cognitive** responses, regardless of ethnicity.<br>- The changes could **not be attributed** to the investigated **mediators**.<br>- Women should be encouraged to go outdoors and specifically visit parks to | - **Environment type** (F/B) - Arab woman demonstrated improvement in most outdoor environments, while for the Jewish woman, improvement was reported mainly in parks, but not in any other urban environment.<br>- **Socio-demographic** (ethnic preferences) (F) - whereby benefits were stronger in intra-ethnic parks.<br>- **Comfort level at home** (F/B) - more comfort at home for Jewish women vs |

| | | | | | | | | |
|---|---|---|---|---|---|---|---|---|
| | | | | | | | improve their psychological and physiological health | Arab women, which could have reduced positive effects of outdoor environments considered less comfortable for this subgroup. |
