## Supplementary Table 4 for "Enabling Health Outcomes of Nature-based Interventions: A Systematic Scoping Review"

| Mental Health Outcomes | | | | |
|---|---|---|---|---|
| *Category* | *Outcome* | *Reference* | *Association (with NOE)* | *Modulators (Barrier/Facilitator)* |
| **Psychological health** | HRQoL – health-related quality of life | [131] [142] | Positive | Stressful life events (B), age (B/F), environment type (F) (131); no difference between intervention vs control (142) |
| | Quality of Life (QoL) | [147][164] | Positive | Changes to the built environment (improved footpaths and clearing of rubbish and vandalism) (F) (147); Environment type (F) (164) |
| | Wellbeing | [132][136] [141][146] [149][154] [164][165] | Positive/ Mixed effects | **Positive**: Environment type (F) (132); environment type (F), previous exposure to nature as child and last year (F), positive affect (F) only in green space, nature connectedness (F) in both control/ intervention (136); physical activity (F), presence of others (B), stressful life events (B) (141); Interpersonal processes (F), Environmental processes (B/F – based on access, perceived aesthetics, and neighbourhood attachment) (146); Environment type (F) (164); Transportation (F), Staff knowledge (F), Group organisation (F), Biodiversity (F) (165) Air and heat-related pollution (B), Proximity (F), SES (B/F) (154);<br>**Mixed effects**: study design (B); terminology for GS (B) (149); |

| | | | | |
|---|---|---|---|---|
| **Psychological health** | Hedonic Wellbeing (life satisfaction) | [149] | Positive | Connectedness with nature (F), Active engagement with nature (F) (149) |
| | Eudaimonic wellbeing (personal flourishing) | [149] | No effect | n/a |
| | Perceived wellbeing | [134][151] [152][153] [161][168] | Positive | Environment type (F), physical activity (F) (134); Environment type (F), Active engagement with nature (F) Safety concerns (B), Staff attitudes and lack of staff education and awareness (B), Social prejudice and stigma (B), Limited staff and resources (B); weather (B/F); Negative self-perception and lack of confidence (B); Poor physical and visual access (B), Poor garden design (B) (i.e. benches, weather protection), Care culture NOT person-centred (B) (151); Micro-features (i.e. benches) (F) (152); Positive staff attitudes (153); Environment type (ocean > public pools) (F) (161); Access (B/F), Environment type (F), Fear and stigma (B), Lack of resources/ equipment (B) (168) |
| | Perceived mental health | [135] | Positive | Safety (B), accessibility (B/F), personal perceptions (B/F) (135) |
| | Depression | [141][142] [156][158] | Negative | Stressful life events (B), physical activity (F) (141); no difference between intervention vs control (142); Presence of caregivers (156); |

| | | | | |
|---|---|---|---|---|
| | | | | Physical activity (F), Environment type (F) (158) |
| **Psychological health** | Anxiety | [142][143] [156][158] [159][165] | Negative | No differences between intervention vs control (142); Season - Summer (F) (143); Presence of caregivers (156); Physical activity (F), Environment type (F) (158); Environment type (forests>roadside> daily activities) (F), Physical activity (F) (159); Transportation (F), Staff knowledge (F), Group organisation (F), Biodiversity (F) (165) |
| | Psychological Restoration | [163] | Positive | Biodiversity in urban park (F) (163) |
| **Social health** | Social isolation | [134][151] [152][153] [161][165] | Negative | Environment type (F); physical activity (F) (134); Environment type (F), Active engagement with nature (F) Safety concerns (B), Staff attitudes and lack of staff education and awareness (B), Social prejudice and stigma (B), Limited staff and resources (B); weather (B/F); Negative self-perception and lack of confidence (B); Poor physical and visual access (B), Poor garden design (B) (i.e. benches, weather protection), Care culture NOT person-centred (B) (151); Micro-features of the environment (benches) (F), Accessibility (B) (152); Positive staff attitudes (F), Presence of others (F) (153); Group membership (F), Environment type (F) (161); Transportation |

| | | | | |
|---|---|---|---|---|
| | | | | (F), Staff knowledge (F), Group organisation (F), Biodiversity (F) (165) |
| **Social health** | Social connectedness | [144][146] [156][160] [161][162] [168] | Positive | Environment type (F), Presence of animals (F) (144); Intrapersonal processes (F), Interpersonal processes (F), Environmental processes (B/F – based on access, perceived aesthetics, and neighbourhood attachment) (146); Environment type, i.e. farms (F), Harvest speed (F) (156); Lower SES (F), Environment type (F) (160); Group membership (F), Weather (F), Threatening biodiversity (F) (161); Environment type (F), Individual factors (i.e. time pressures, changing identities) (B), Presence of others (B/F) (162); Access (B/F), Fear and stigma (B), Lack of resources/ equipment (B), Environment type (F) (168) |
| | Social discomfort | [169] | Negative | Environment type (F/B), Ethnicity (B/F) (169) |
| **Emotional Health** | Positive Affect | [136][142] [148][151] [159][160] [161] [164] [165] | Positive | Environment type (F) (136); Environment type (F), physical activity (F), stressful life events (B) (141); Environment type (forests>roadside> daily activities) (F), Physical activity (F) (159); Environment type (ocean>pool) (F), Biodiversity – if non-threatening (F) (161); Environment type (natural>control) (F) (164) Micro-features (F), Conditions of natural environments (B/F), Perceived quality (F), Accessibility |

| | | | | |
|---|---|---|---|---|
| **Emotional Health** | | | | (B/F), Safety (B) (148); Environment type (F), Active engagement with nature (F) Safety concerns (B), Staff attitudes and lack of staff education and awareness (B), Social prejudice and stigma (B), Limited staff and resources (B); weather (B/F); Negative self-perception and lack of confidence (B); Poor physical and visual access (B), Poor garden design (B) (i.e. benches, weather protection), Care culture NOT person-centred (B) (151); Environment type (F), Physical activity (F) (160); Transportation (F), Staff knowledge (F), Group organisation (F), Biodiversity (F) (165) |
| | Positive mood state | [140][143] [157][158] [167][169] | Positive | Environment type (F) (140); Physical activity (F), Environment type (F) (158); Conservation (F), Physical activity (F), Social interaction (F) (167) |
| | Negative affect | [141][159] [161][164] | Negative | Stressful life events (B), physical activity (F) (141); Environment type (forests>roadside> daily activities) (F), Physical activity (F) (159); Environment type (ocean>pool) (F), Biodiversity (threatening) (B) (161); Environment type (F) (164) |
| | Mood disturbance | [143] | Negative | Seasons – Spring (F) (143) |
| | Self-esteem | [139][151] [158][162] [168] | Positive | Gender – more effect for women vs men (F), Duration of intervention (F) (139); Environment type (F), Active engagement with nature (F), Safety concerns (B), Staff |

| | | | | |
|---|---|---|---|---|
| **Emotional Health** | | | | attitudes and lack of staff education and awareness (B), Social prejudice and stigma (B), Limited staff and resources (B); weather (B/F); Negative self-perception and lack of confidence (B); Poor physical and visual access (B), Poor garden design (B) (i.e. benches, weather protection), Care culture NOT person-centred (B) (151); Physical activity (F), Environment type (F) (158); Environment type (F), Individual factors (i.e. time pressures, changing identities) (B), Presence of others (B/F) (162); Access (B/F), Fear and stigma (B), Lack of resources/equipment (B) (168) |
| | Self-confidence | [153] | Positive | Active engagement in nature (F), Presence of others (F) (153) |
| | Vitality | [140][144] | Positive | Environment type (F) (140); Environment type (F), Presence of animals (F) (144) |
| | Agitation | [151][156] | Negative | Environment type (F), Active engagement with nature (F) (151); Garden design (F), Mobility (F), Activity itself (TH) (F) (156) |
| **Emotional Health** | Behavioural Problems (i.e. inattention, hyperactivity, violence) | [150][151] [157[[158] | Negative | Physical activity (F), Environment type (F) (150); Environment type (F), Active engagement with nature (F) (151); Environment type (F), Accessibility (B), Ethnicity, i.e. south Asian children (B), Deprivation (B), Quality of GS (B/F) (158); Environment type (F), Physical activity (F) (157) |

| | | | | |
|---|---|---|---|---|
| | Perceived restoration | [137][138] [140][143] [160][169] | Positive | Environment type (F) < perceived sound quality (F) (137); sensory qualities (F), safety (B), topography (F) (138); Environment type (F) (140); presence of water (F) (143); Environment type (F) (160); Environment type (F) (169) |
| **Stress** | Perceived stress | [141][164] [165] | Negative | Stressful life events (B), physical activity (F) (141); Environment type (F) (164); Transportation (F), Staff knowledge (F), Group organisation (F), Biodiversity (F) (165) |
| | Psychological resistance | [132][134] [168] | Positive | Environment type (F) (132); Environment type (F) (134); Access (B/F), Fear and stigma (B), Lack of resources/equipment (B) (168) |
| **Stress** | Stress reduction | [132][141] [144][151] [160][159] [161][162] [167] | Positive | Environment type (F) (132); Presence of others (B), Physical activity (F) (141); Environment type (F), Presence of animals (F) (144); Environment type (F), Active engagement with nature (F) Safety concerns (B), Staff attitudes and lack of staff education and awareness (B), Social prejudice and stigma (B), Limited staff and resources (B); weather (B/F); Negative self-perception and lack of confidence (B); Poor physical and visual access (B), Poor garden design (B) (i.e. benches, weather protection), Care culture NOT person-centred (B) (151); Environment type (forests>roadside> daily activities) (F), |

| | | | | |
|---|---|---|---|---|
| | | | | Physical activity (F) (159); Environment type (F) (160); Activity itself – i.e. swimming in ocean (F) (161); Environment type (i.e. trees, plants, views, etc.) (F), Poor quality of GS/BS (B), Deprivation (B) (162); Conservation (F), Physical activity (F), Social interaction (F), Location of NBI (natural reserve>bog>field) (F) (167) |
| | Psychological distress | [158] | Negative | Environment type (F), Accessibility (B), Ethnicity, i.e. south Asian children (B), Deprivation (B) (158) |
| **Physiological Outcomes** | | | | |
| *Category* | *Outcome* | *Reference* | *Association (with NOE)* | *Modulators (Barrier/Facilitator)* |
| **Cardio-vascular outcomes** | Blood pressure (systolic and diastolic) | [132][140] [143][145] | Negative/ No effects | **NEGATIVE**: Environment type (F) (132); Environment type (F), activity itself (viewing>walking) (F) (140); Environment type (urban park vs control), seasons (summer) (F/B) (143); **NO EFFECTS**: temperature (B), humidity (B), light spectrum (G/B ratio too high) (B) (145) |
| | Heart rate | [132][140][ 143] [145] [157] | Negative/ Positive | **Negative**: Environment type (F) (132); Environment type (F) – in favour of urban forests > urban parks, noise pollution (B), air pollution (B) (140) Features of the environment (trees species – where |

| | | | | |
|---|---|---|---|---|
| | | | | maple>oak>birch) (F) (145); Environment type (green>control) (F), Physical activity (F) (157)<br>**Positive**: environment type (post-viewing nature) (F) (143); |
| | Heart rate variability (HRV) - SDNN | [140]<br>[157][169] | Positive/<br>mixed effects | **Positive**: Environment type (F) – increased in green environment vs control (140); Environment type (F) (parks>urban) (169)<br>**No effects**: Environment type (F), Poor study design (B) (157) |
| **Stress** | Cortisol | [140][160] | Negative | Environment type (F) – all decrease but green (forests>park) > control (140); Environment type (F) (160) |
| **Physical Health Outcomes** | | | | |
| *Category* | *Outcome* | *Reference* | *Association (with NOE or NBI)* | *Modulators (Barrier/Facilitator)* |
| | Physical activity (LTPA) | [133] | Positive | Safety concerns (B), presence of others (B/F), accessibility (F), natural environment (F), environmental design (B/F) (133) |
| | Physical activity – swimming | [134] | Positive | Fear and stigma on body type (B) (134) |
| | Physical activity – walking in nature | [138][154] | Positive | Proximity (F) (138); Air and heat-related pollution (B), Proximity (F), SES (B/F) (154) |

| | | | | |
|---|---|---|---|---|
| **Physical activity in GS or BS** | Perceived physical health | [134][135] [146] | Positive | Physical activity in sea (F) (134); Safety (B), accessibility (B/F), personal perceptions (B/F) (135); Interpersonal processes (F), Environmental processes (B/F – based on access, perceived aesthetics, and neighbourhood attachment) (146) |
| | Physical activity in Urban Green Spaces (UGS) | [147][152] [160] | Positive | Changes to the built environment in parks (F), Proximity to newly built cycling/walk lanes (F) (147); Micro-features of the environment (benches) (F), Gender (F – for sedentary women), Accessibility (B) (152); Accessibility (B), Attractivity and activity in programs (B/F), Age (F) – i.e. children and young adults (160) |
| | Physical activity (MVPA) | [157][166] | Positive | Exposure to GS (F) (157); Duration, i.e. longer time in GS (F), Environment type (F) (166) |
| **Physical activity in GS or BS** | Sedentary Time | [166] | Negative | Duration, i.e. longer time in GS (F), Environment type (F) (**166**) |
| | Physical fitness | [168] | Positive | Environment type (F), Intervention (i.e. surfing) (F), Access (B/F), Fear and stigma (B), Lack of resources/equipment (B) (168) |
| **Fatigue** | Post-stroke fatigue (PSF) | [142] | Negative | Intervention (F) (142) |
| **Mortality** | All-cause mortality | [148] [157] | Negative | Positive affect (F), Heat reduction (F), Environment type (F) (148); Environment type (F) (157) |
| **General physical health** | Overall health | [150][158] [165] | Positive | Physical activity (F), Environment type (F), Accessibility (F), Quality and quantity of GS (F) (150); Physical activity (F), Environment |

| | | | | |
|---|---|---|---|---|
| | | | | type (F) (158); Transportation (F), Staff knowledge (F), Group organisation (F), Biodiversity (F) (165) |
| **Motor functioning** | Mobility | [152] | Positive | Micro-features of the environment, i.e. benches (F), Injury (B), Engagement in social interactions (F), Accessibility (B) (152) |
| | Disability | [142] | Negative | No differences between intervention vs control – both decreased (142) |
| **Recovery** | Recovery (from mental illnesses) | [153] | Positive | Presence of others (F), Active engagement in nature (F) (153) |
| **Obesity** | Obesity | [148][154] [160][165] | No effect | Quality of study design (B) (148, 154, 160) |
| **Sleep** | Sleep (quality and quantity) | [155] | Positive | Time of day (afternoon>morning for walking) (F), Environment type (outdoors>indoors) (F), Behavioural contexts (weekdays vs weekends preferences for different GS) (F) (155) |
| **Cognitive Outcomes** | | | | |
| *Category* | *Outcome* | *Reference* | *Association (with NOE)* | *Modulators (Barrier/Facilitator)* |
| | Science, technology, engineering, and math (STEM)-capacity | [131] | Positive | Duration of intervention (F); stressful life events (B); environment type (F) (131) |

| | | | | |
|---|---|---|---|---|
| **Cognition** | Attention Retention | [137][144] [157][160] | No effect/ Positive | **No effects** poor quality of measurements (B) (137); **Positive**: Environment type (F), Presence of animals (F) (144); Environment type (F), Physical activity (F) (157); Poor study design (B), Environment type (F) (160) |
| **Cognition** | Attention restoration | [132][150] | Positive | Environment type (F) (132); Physical activity (F), Proximity (F), Environment type (F), Accessibility (F), Quality and quantity of GS (F) (150) |
| **Symptom reduction** | ADHD symptoms | [150] | Negative | Attention restoration (F), Spatial working memory (F), environment type (F) (150) |
| **Memory** | Spatial working memory | [150][169] | Positive/ no effects | **Positive**: Physical activity (F), Proximity (F), Environment type (F) (150); **No effects**: (169) |
| | Executive functioning (I.e. memory) | [156][159] | Positive/ no effects | **Positive**: Active engagement in activity (F) (156) **No effects**: Poor study design (B), Poor measurements (B) (159) |
